# Genetic architecture of 67 oral diseases and their links to systemic diseases

**DOI:** 10.1101/2025.10.16.25338184

**Authors:** Kirika Karppinen, Hanna M Ollila, Kanwal Batool, FinnGen, Estonian Biobank Research Team, Erik Abner, David P Rice, Aarno Palotie, Tuula Palotie, Samuli Ripatti, Nina Mars, Satu Strausz

## Abstract

Oral and craniofacial diseases are common, yet their genetic basis and links to systemic health are incompletely understood. We performed genome-wide association analyses of 67 oral phenotypes in 500,348 FinnGen participants, identifying 105 genome-wide significant loci, including 47 previously unreported associations. Fine-mapping revealed 14 coding variants, such as a missense variant in *USP31* for caries and in *MANBA* for oral leukoplakia, and a stop-gained variant in *GPNMB* for temporomandibular disorders. HLA analyses implicated *DQA1* and *DQB1* alleles in lichen planus and other mucosal disorders. We observed 378 statistically significant genetic correlations among oral traits, such as tooth loss and chronic apical periodontitis (*r_g_* = 0.91, 95% CI [0.76 – 1.05], *P* = 1.73 × 10^-34^), and 419 significant correlations between oral and systemic diseases, including periodontal diseases with chronic laryngitis (r_g_ = 0.97, CI [0.58 – 1.36], *P* = 1.23 × 10^-6^) and bruxism with gastro-oesophageal reflux (r_g_ = 0.51, CI [0.38 – 0.65], *P* = 1.10 × 10^-13^). These results expand the catalog of oral disease loci, uncover Finnish-enriched risk alleles, and highlight shared inflammatory, immune, and structural pathways connecting oral and systemic health.

## Introduction

Oral diseases, including dental caries, periodontitis and temporomandibular disorders (TMD), contribute significantly to global disease burden, with untreated dental caries affecting 2.3 billion people and severe periodontitis affecting over 750 million individuals globally [1]. Despite epidemiologically well-established connections to systemic conditions, including cardiovascular disease, diabetes, and autoimmune disorders [2–4], the underlying biological and genetic factors remain incompletely characterized. Previous genome-wide association studies have reported SNP-based heritability of 13 % for dental caries and 1-6 % for periodontitis depending on phenotype definition [5]. Large-scale cohorts with genetic information and diagnostic codes provide an opportunity to explore the shared genetic architecture across different oral diseases, and their overlap with systemic diseases.

Oral diseases are complex, multifactorial conditions influenced by genetic, environmental, inflammatory, and microbiome-related factors. While genome-wide association studies (GWAS) have primarily focused on common conditions such as caries and periodontitis [5–8], recent efforts have begun to investigate a broader range of oral phenotypes, including pulp and periapical diseases, bruxism, and dentofacial developmental anomalies [9–13]. These studies have discovered risk loci involved in immune regulation and barrier defense (e.g. *SIGLEC5*, *DEFA1A3*), craniofacial development (e.g. *IRF6*), and HLA-mediated immune pathways, highlighting the links of oral traits to systemic health [6, 11–13]. However, most GWASs to date remain limited in scale or scope, focusing often on single diseases. This leaves the broader genetic landscape of oral health and the overlap with systemic diseases largely uncharacterized.

We therefore aimed to conduct a comprehensive genome-wide investigation of 67 common and rare oral and craniofacial phenotypes within the FinnGen study [14], leveraging germline genetic data linked to nationwide health registries covering both systemic and dental diagnoses in 500,348 individuals. Here, we explore the unique and shared genetic architecture of oral and craniofacial phenotypes and their links to systemic diseases. Our study discovers new loci associated with oral and craniofacial phenotypes and provides insights into genetic risk factors that link oral diseases and general health.

## Results

### Oral phenotypes span a wide clinical spectrum

The FinnGen dataset includes 500,348 individuals with genome-wide genotyping and comprehensive health registry data, including lifetime medical diagnoses, prescription and purchase data, laboratory values, sociodemographic factors, and cause-of-death records. We assessed 67 oral and craniofacial phenotypes that span a range of structures from dental hard tissue to oral mucosa and salivary glands (Fig. 1a, Supplementary Table 1). All 67 phenotypes were grouped into seven categories based on tissue type and diagnostic similarity, as visualized in Fig. 1b and 1c.

**Figure 1.**
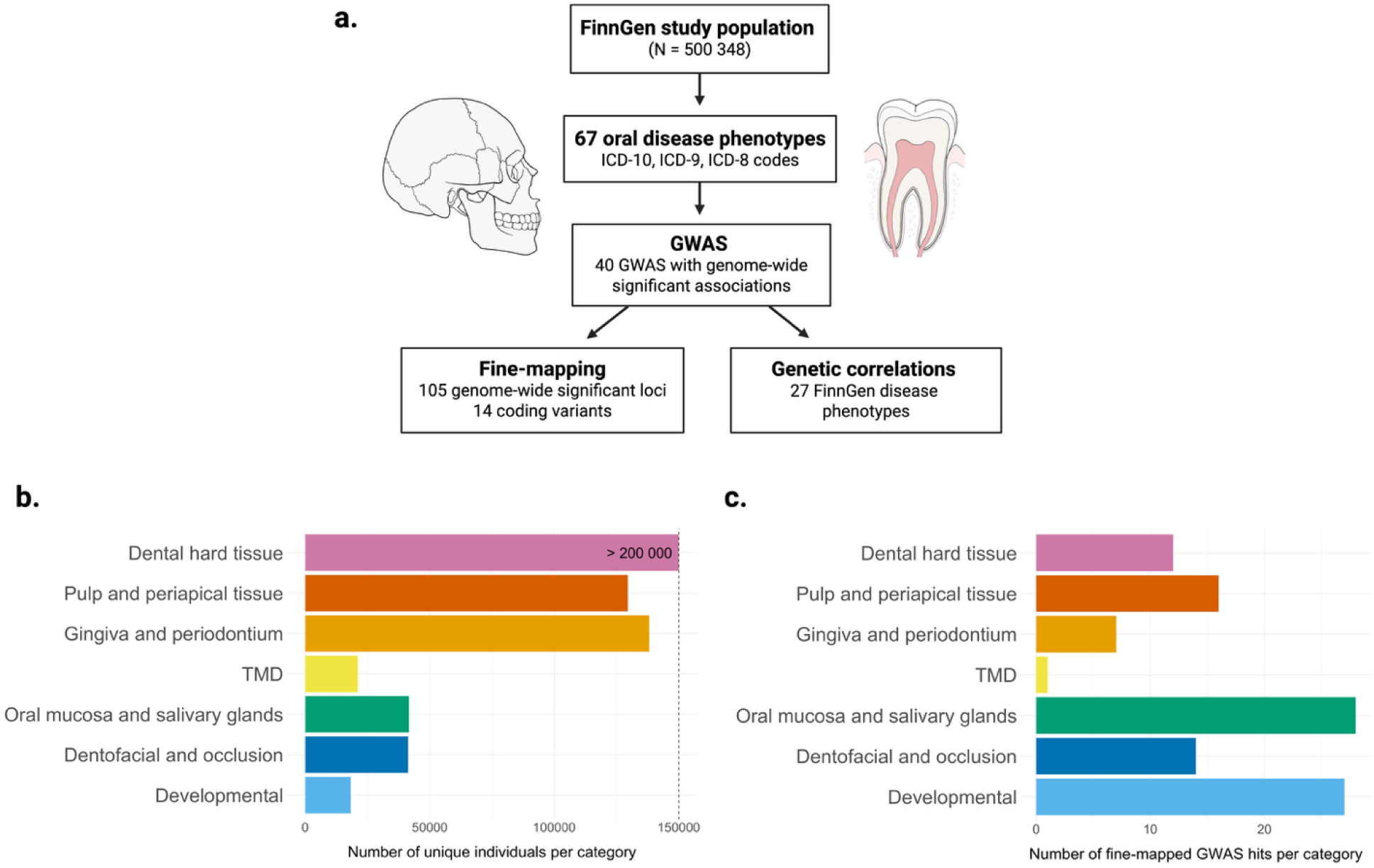
Study overview. a) Flowchart of study design and data. b) Number of unique cases per phenotype category. 67 oral and craniofacial phenotypes are grouped into 7 phenotype categories. In this visualization, cases are included only once in each category. The x-axis has been limited to 200 000 for visualization purposes. c) Number of fine-mapped GWAS hits per phenotype category. Here, loci shared between several phenotypes are included only once in each category.

The most prevalent diseases were dental caries (N = 223,126) and gingivitis and periodontal diseases (N = 137,746), followed by diseases of pulp and periapical tissues (N = 129,318), chronic apical periodontitis (N = 75,784), and pulpitis (N = 50,909). Rarer diagnoses included oral leukoplakia (N = 930), cleft palate (N = 388), and impacted maxillary canine (N = 311), highlighting the wide phenotypic range of the dataset (Fig. 1b, Supplementary Table 1, Supplementary Figure 1).

### Genome-wide association analysis discovers 105 variants for oral and craniofacial phenotypes

We conducted GWASs for all 67 phenotypes and identified genome-wide significant associations for 40 phenotypes [15]. For these 40 phenotypes, we performed fine-mapping [16] to identify lead and credible set variants, which revealed 105 independent genome-wide significant loci (Supplementary Table 2). We discovered lead variants for high-burden traits such as caries (e.g., rs187591243 in *LASP1*, *β* = 0.14, *P* = 7.9 × 10^-9^) and gingivitis/periodontitis (e.g., rs187684552 in *SIPA1L1*, *β* = 0.24, *P* = 1.2 × 10^-8^), as well as for developmental anomalies such as cleft hard palate (rs187395934 in *RPS6KA1*, *β* = 3.46, *P* = 4.9 × 10^-8^) and impacted maxillary canine (rs189229384 in *PABPC4L*, *β* = 1.34, *P* = 3.1 × 10^-8^). A subset of variants showed notably large effect sizes, particularly those associated with rare traits like hypertrophy of tongue papillae (rs1178338740 in *CMKLR2, β* = 3.2, *P* = 1.6 × 10^-8^) and maxillary hypoplasia (rs573825482 in *TMEM92, β* = 4.7, *P* = 8.9 × 10^-9^) (Table 1).

**Table 1.** Lead variants of previously unreported loci associated with oral phenotypes in FinnGen. Genome-wide association analysis identified 105 loci including previously unreported loci across 67 oral and craniofacial phenotypes. Variants shown are the most strongly associated in each locus according to fine-mapping, and the loci have no previously reported genome-wide significant associations with the same phenotype category in the GWAS Catalog [17]. Chrom=chromosome, pos=position, ref=reference allele, alt=alternative allele, β=beta, se=standard error, p=p-value, PIP=posterior inclusion probability, af = allele frequency in FinnGen for the alternative allele, FIN=Finnish enrichment of variants found in gnomAD 3.1.2. Variants not found in gnomAD are marked with NA.

| phenotype | rsid | chrom:pos:ref:alt | $\beta$ | se | p | PIP | af | FIN | variant | gene |
| --- | --- | --- | --- | --- | --- | --- | --- | --- | --- | --- |
| Attrition | rs42131 | 7:73397257:A:T | -0.070 | 0.012 | 7.50e-09 | 0.491 | 0.195 | 1.084 | regulatory region variant | FZD9 |
| Tooth wear | rs188833787 | 3:76017712:A:T | 0.224 | 0.041 | 4.03e-08 | 0.471 | 0.008 | 7.149 | intron variant | ROBO2 |
| Tooth wear | rs42129 | 7:73408518:C:A | -0.061 | 0.010 | 9.15e-10 | 0.459 | 0.176 | 1.054 | upstream gene variant | FZD9 |
| Tooth wear | rs2093587330 | 11:3989373:A:G | 0.817 | 0.149 | 4.63e-08 | 0.439 | 0.001 | NA | noncoding transcript exon variant | STIM1 |
| Caries | rs200486134 | 16:23069106:C:G | 0.864 | 0.149 | 7.35e-09 | 0.569 | <0.000 | 0.473 | missense variant | USP31 |
| Caries | rs187591243 | 17:38913563:G:A | 0.141 | 0.024 | 7.92e-09 | 0.360 | 0.007 | 1.726 | intron variant | LASP1 |
| Diseases of pulp and periapical tissues | rs4988521 | 2:229754017:C:G | -0.757 | 0.137 | 3.12e-08 | 0.655 | <0.000 | 0.157 | intergenic variant | TRIP12 |
| Diseases of pulp and periapical tissues | rs560704998, rs75187107, rs398106121 | 3:85546839:C:CA | -0.028 | 0.005 | 3.77e-08 | 0.005 | 0.711 | 1.164 | intron variant | CADM2 |
| Diseases of pulp and periapical tissues | rs5871429, rs397804460 | 5:131585275:G:GA | 0.026 | 0.005 | 4.80e-08 | 0.012 | 0.619 | 0.865 | intron variant | RAPGEF6 |
| Pulpitis | rs1321699072 | 10:89325978:C:T | -0.629 | 0.115 | 4.95e-08 | 0.651 | 0.002 | 18.719 | intron variant | LIPA |
| Chronic apical periodontitis | rs34648990 | 20:52084522:G:A | 0.316 | 0.058 | 4.51e-08 | 0.586 | 0.002 | 0.101 | 3 prime UTR variant | ZFP64 |
| Gingivitis and periodontal diseases | rs7613444 | 3:52504284:G:C | 0.030 | 0.005 | 2.91e-08 | 0.866 | 0.236 | 1.471 | intron variant | STAB1 |
| Gingivitis and periodontal diseases | rs111231814 | 12:24820195:C:A | -0.630 | 0.108 | 4.94e-09 | 0.446 | 0.001 | 0.52 | intron variant | BCAT1 |
| Gingivitis and periodontal diseases | rs187684552 | 14:71584969:A:G | 0.244 | 0.043 | 1.19e-08 | 0.643 | 0.003 | 0.268 | intron variant | SIPA1L1 |
| Chronic complicated periodontitis | rs75351459 | 3:141093823:C:T | 0.109 | 0.020 | 3.76e-08 | 0.564 | 0.059 | 1.997 | intron variant | SPSB4 |
| Chronic complicated periodontitis | rs2877160 | 5:30781297:C:T | 0.058 | 0.010 | 7.23e-09 | 0.880 | 0.62 | 0.895 | intergenic variant | CDH6 |
| Chronic complicated periodontitis | rs1051730 | 15:78601997:G:A | 0.056 | 0.010 | 2.38e-08 | 0.070 | 0.331 | 0.914 | synonymous variant | CHRNA3 |
| Embedded and impacted teeth | rs1182052384 | 3:138799892:T:C | 0.582 | 0.102 | 1.15e-08 | 0.992 | 0.003 | Inf | intron variant | PIK3CB |
| Impacted maxillary canine | rs189229384 | 4:135451123:A:T | 1.344 | 0.243 | 3.09e-08 | 0.175 | 0.011 | 1.709 | intergenic variant | PABPC4L |
| Enamel hypoplasia | rs117645102 | 18:52010029:G:A | 2.469 | 0.443 | 2.48e-08 | 0.737 | 0.001 | 7.794 | intergenic variant | DCC |
| Hypodontia | rs531152356 | 6:5323510:T:G | 2.395 | 0.439 | 4.91e-08 | 0.412 | <0.000 | 0 | intron variant | FARS2 |
| Hypodontia | rs2320968 | 13:22218835:A:G | 0.285 | 0.049 | 5.93e-09 | 0.573 | 0.146 | 1.072 | intron variant | FGF9 |
| Cleft hard palate | rs187395934 | 1:26589269:G:A | 3.462 | 0.635 | 4.86e-08 | 0.951 | 0.001 | NA | downstream gene variant | RPS6KA1 |
| Anomalies in dental arch relations | rs149999594 | 10:35697922:C:T | -0.406 | 0.070 | 7.61e-09 | 0.915 | 0.013 | 1.23 | noncoding transcript exon variant | FZD8 |
| Anomalies in dental arch relations | rs1443684783 | 18:77083370:TC:T | 1.102 | 0.191 | 8.02e-09 | 0.957 | 0.001 | Inf | intron variant | MBP |
| Asymmetry of jaw | rs1412908547 | 17:6086430:G:C | 3.738 | 0.652 | 1.01e-08 | 0.993 | <0.000 | NA | intron variant | WSCD1 |
| Crowding of teeth | rs201475998 | 2:108875328:AAAAAAAAG:A | 0.260 | 0.046 | 1.25e-08 | 0.253 | 0.055 | 8.59 | intron variant | CCDC138 |
| Crowding of teeth | rs929805940 | 3:2153482:T:C | 1.879 | 0.339 | 3.00e-08 | 0.974 | 0.001 | NA | intron variant | CNTN4 |
| Deep bite | rs4938326 | 11:117034748:G:C | 0.172 | 0.031 | 3.53e-08 | 0.107 | 0.114 | 1.268 | intron variant | SIK3 |
| Deep bite | rs1257741 | 12:28381636:G:C | -0.144 | 0.025 | 1.11e-08 | 0.078 | 0.227 | 0.906 | intron variant | CCDC91 |
| Maxillary hypoplasia | rs573825482 | 17:50249671:G:A | 4.720 | 0.821 | 8.89e-09 | 0.988 | <0.000 | 8.049 | intergenic variant | TMEM92 |
| Mandibular prognathia | rs535975844 | 13:96784159:T:G | 2.543 | 0.455 | 2.30e-08 | 0.421 | 0.001 | Inf | intron variant | HS6ST3 |
| Mandibular retrognathia | rs227727 | 17:56699594:A:T | 0.151 | 0.027 | 3.89e-08 | 0.286 | 0.487 | 1.124 | intergenic variant | NOG1 |
| Mandibular retrognathia | rs7225448 | 17:71385926:A:G | 0.153 | 0.028 | 2.96e-08 | 0.138 | 0.433 | 1.169 | intergenic variant | SOX9 |
| Open bite | rs139374150 | 3:10913763:G:A | 0.685 | 0.126 | 4.93e-08 | 0.962 | 0.011 | 2.105 | intron variant | SLC6A11 |
| Open bite and surgery | rs572104116 | 12:101645832:G:A | 3.019 | 0.533 | 1.47e-08 | 0.507 | 0.001 | 4.689 | intron variant | MYBPC1 |
| Maxillary retrognathia | rs11073419 | 15:95119863:T:C | -0.396 | 0.072 | 3.09e-08 | 0.888 | 0.206 | 0.882 | intron variant | MCTP2 |
| Temporomandibular disorders | rs191297708 | 7:23217214:T:C | -0.125 | 0.021 | 1.99e-09 | 0.047 | 0.067 | 18.635 | intergenic variant | NUP42 |
| Oral cysts | rs181890726 | 7:63794711:A:G | -0.539 | 0.097 | 3.05e-08 | 0.141 | 0.035 | 0.677 | downstream gene variant | ZNF722 |
| Stomatitis | rs201138667 | 4:96283898:TA:T | 0.424 | 0.077 | 4.46e-08 | 0.331 | 0.009 | NA | intergenic variant | PDHA2 |
| Oral leukoplakia and other epithelial disorders | rs74649783 | 4:102502645:A:G | 0.444 | 0.071 | 3.19e-10 | 0.607 | 0.024 | 1.848 | intron variant | NFKB1 |
| Oral leukoplakia and other epithelial disorders | rs542270346 | 6:110242809:T:C | 0.987 | 0.170 | 6.61e-09 | 0.936 | 0.004 | 1.366 | intron variant | CDC40 |
| Oral leukoplakia | rs755173701 | 2:234643265:T:C | 2.385 | 0.434 | 4.03e-08 | 0.573 | 0.001 | 3.122 | intergenic variant | ARL4C |
| Oral leukoplakia | rs1172595265 | 6:22745355:G:A | 4.742 | 0.775 | 9.17e-10 | 0.565 | <0.000 | 0 | intron variant | HDGFL1 |
| Peripheral and oral lichen planus | rs189942924 | 6:23106848:T:C | 0.557 | 0.099 | 1.82e-08 | 0.476 | 0.005 | 56.253 | downstream gene variant | HDGFL1 |
| Diseases of salivary glands | rs760512017 | 8:117300695:C:CAAA | 0.614 | 0.106 | 6.47e-09 | 0.993 | 0.003 | 2.675 | regulatory region variant | SLC30A8 |
| Hypertrophy of tongue papillae | rs1178338740 | 2:206245999:AAAAAG:A | 3.205 | 0.567 | 1.63e-08 | 0.417 | <0.000 | NA | intron variant | CMKLR2 |

### Coding variants from fine-mapped loci

Among the lead variants, we discover a previously unreported missense variant for caries (rs200486134 in *USP31*, *β* = 0.86, *P* = 7.4 × 10^-9^). When examining the credible set variants of fine-mapped loci, nine additional coding variants were identified, including five previously unreported variants (Table 2, Supplementary Tables 3-4). Additional coding variants included missense variants for oral leukoplakia and other epithelial disorders (rs75826658 in *MANBA*, *β* = 0.43, *P* = 1.0 × 10^-9^, posterior inclusion probability (PIP) = 0.20) and hypodontia/oligodontia (rs4129190 in *THNSL2*, *β* = −0.32, *P* = 3.5 × 10^-10^, PIP = 0.08), and a stop gained variant for TMD (rs11537976 in *GPNMB*, *β* = −0.13, *P* = 2.2 × 10^-9^, PIP = 0.04). In addition, several known associations were replicated, such as missense variants for hypodontia/oligodontia (rs121908120 in *WNT10A*, *β* = 1.07, *P* = 1.3 × 10^-11^, PIP = 0.48) and orofacial cleft phenotypes (rs41268753 in *GRHL3*).

**Table 2.**
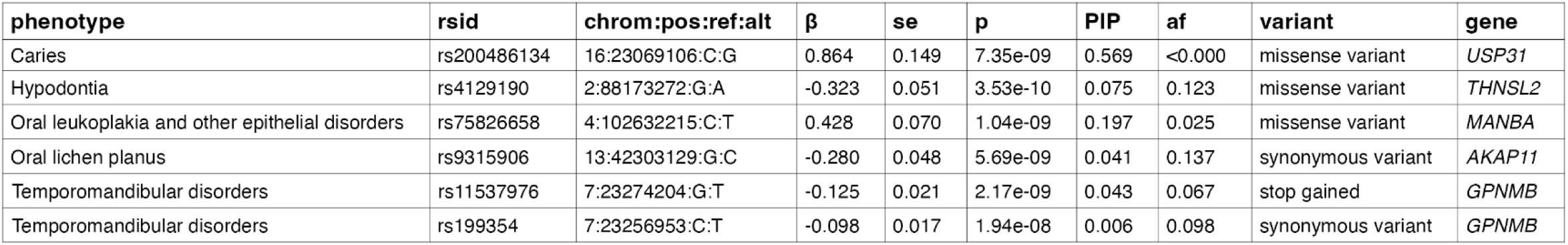
Lead and credible set coding variants associated with oral phenotypes in FinnGen. Genome-wide association analysis identified 14 coding variants from fine-mapped loci associated with oral and craniofacial phenotypes. Variants shown are coding lead variants and coding variants from 95% credible sets, which have no previously reported genome-wide significant associations in the GWAS Catalog [17]. Chrom=chromosome, pos=position, ref=reference allele, alt=alternative allele, β=beta, se=standard error, p=p-value, PIP=posterior inclusion probability, af=allele frequency in FinnGen for the alternative allele.

### 70 variants highlight biology of underexplored traits

To assess the chromosomal architecture of oral disease risk, we mapped the locations of lead variants across the genome. The lead variants were distributed across all autosomes, with notable clusters on chromosomes 1, 2, 4, and 7 (Fig. 2). The genome-wide significant associations span diverse phenotype categories, emphasizing the polygenic nature of oral disease susceptibility.

**Figure 2.**
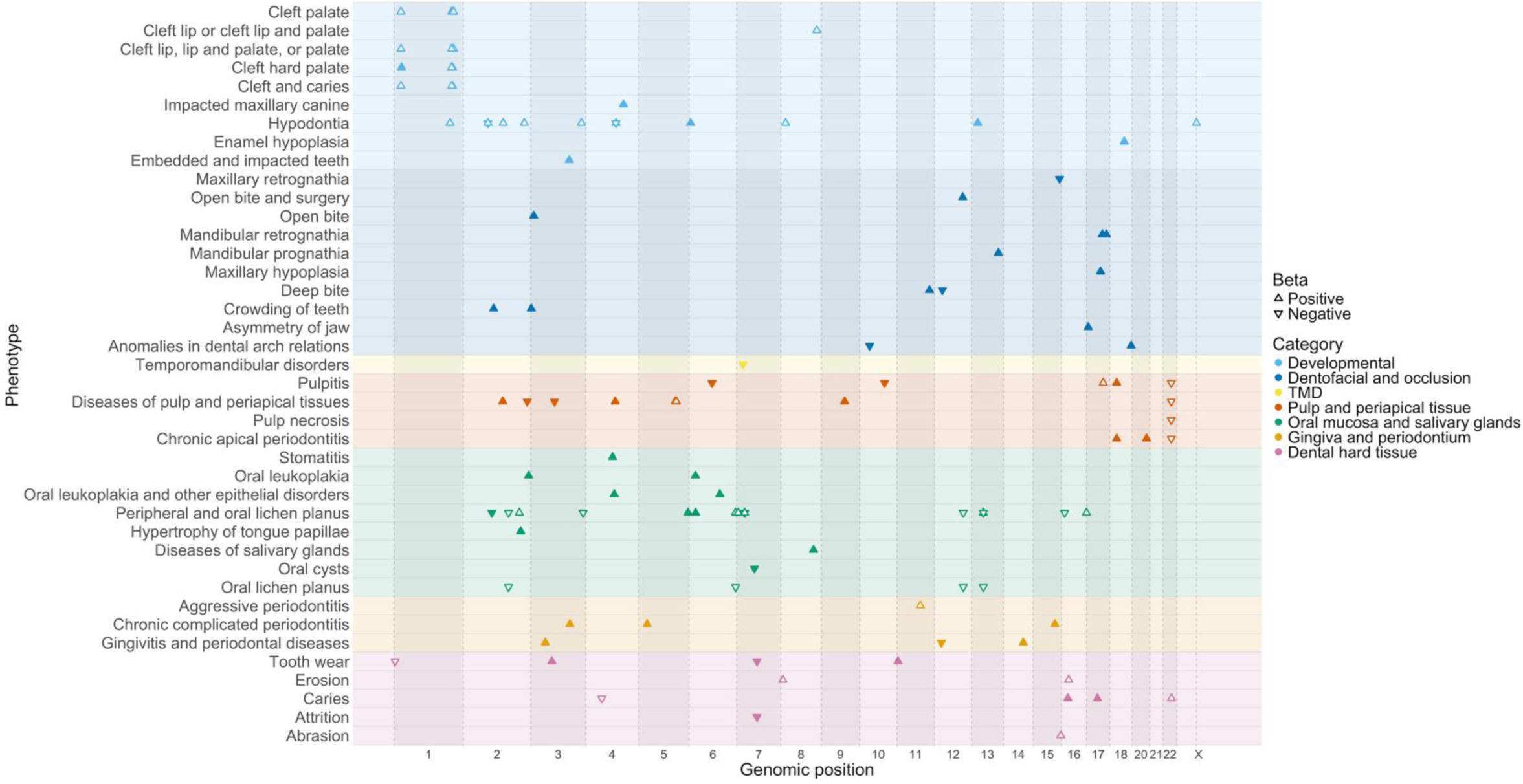
The distribution of fine-mapped GWAS loci across chromosomes for oral and craniofacial phenotypes. The approximate genomic positions of the variants are shown for each phenotype on the x-axis. Phenotypes are organized by phenotype category on the y-axis. Lead variants of previously unreported loci are highlighted. The direction of the triangle indicates the sign of the lead variant beta value.

Of the 105 fine-mapped loci, 47 had no prior associations in previously published GWAS of similar oral phenotypes based on GWAS Catalog and literature search. 70 lead variants had no prior associations in any previously published GWAS. These loci were particularly enriched in dentofacial traits: among the 47 previously unreported loci, one third (14/47) were in dentofacial anomalies and malocclusions, for example, mandibular prognathia (rs535975844 in *HS6ST3*, *β* = 2.54, *P* = 2.3 × 10^-8^) and crowding of teeth (rs929805940 in *CNTN4*, *β* = 1.88, *P* = 3.0 × 10^-8^) (Table 1).

Several of the lead variants were enriched in the Finnish population (FIN), such as rs189942924 in peripheral and oral lichen planus (*HDGFL1*, *β* = 0.56, *P* = 1.8 × 10^-8^, FIN = 56.3), rs1321699072 in pulpitis (*LIPA*, *β* = −0.63, *P* = 5.0 × 10^-8^, FIN = 18.72), and rs191297708 in TMD (*NUP42*, *β* = −0.13, *P* = 2.0 × 10^-9^, FIN = 18.6). In addition, a low-frequency variant rs1443684783 in anomalies in dental arch relations (*MBP*, *β* = 1.10, *P* = 8.0 × 10^-9^, AF = 0.001) was observed only in the Finnish population (Table 1).

### HLA alleles are associated with mucosal and pulpal diseases

We next focused on the Human Leukocyte Antigen (HLA) region to investigate immunogenetic contributions to oral diseases using Finnish specific reference panel for HLA alleles [18].

HLA-fine-mapping revealed strong associations across a range of oral phenotypes, including mucosal, pulpal, and periodontal conditions (Fig. 3, Supplementary Table 5). Most associations, with the exception of attrition, mapped to the HLA class II region, which encodes molecules responsible for presenting extracellularly derived peptides to CD4⁺ T cells, thereby initiating and regulating adaptive immune responses.

**Figure 3.**
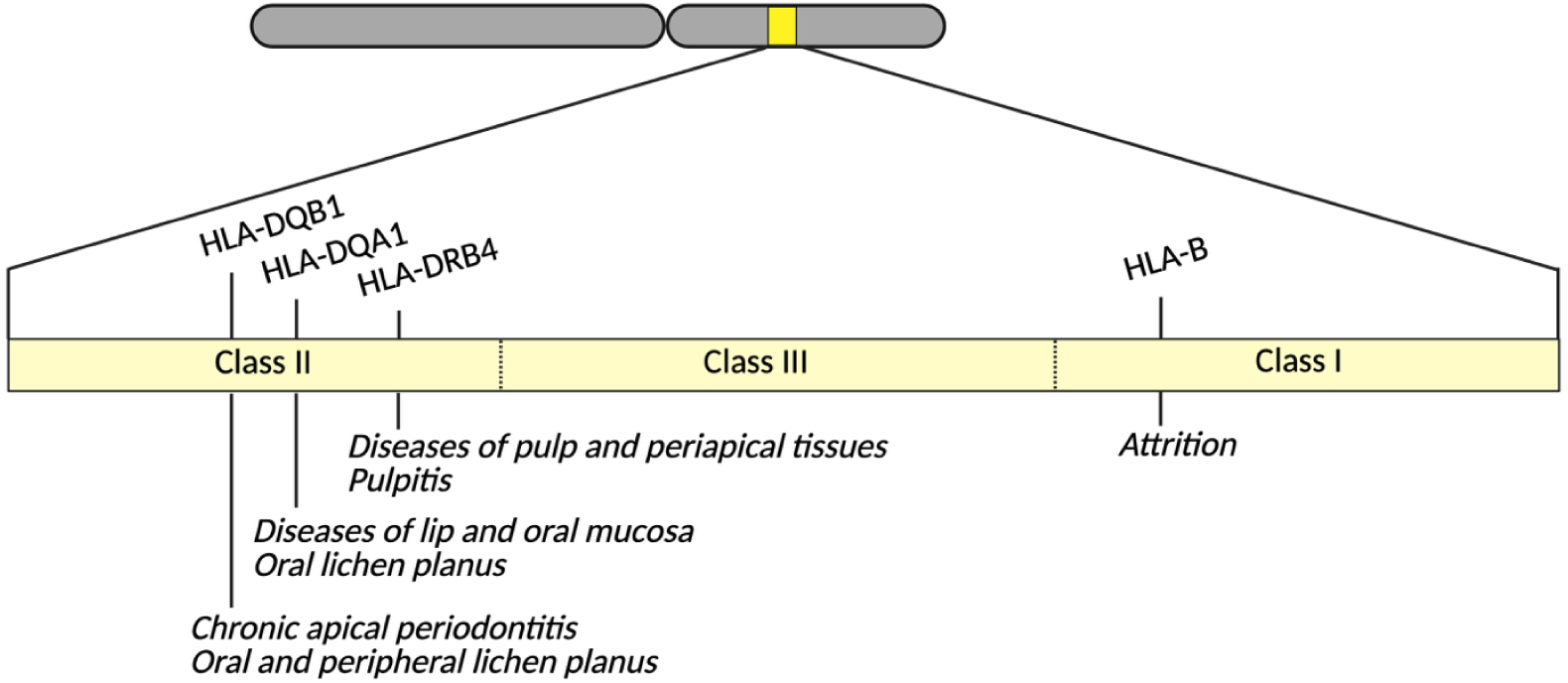
Association of oral phenotypes with HLA alleles according to HLA fine-mapping results. The approximate positions of the associated HLA alleles within the HLA region are shown. The figure is not drawn to scale.

The strongest associations were observed for oral lichen planus (*DQB1*05:01*, *β* = 0.56, *P* = 7.7 × 10⁻²³⁵) and diseases of pulp and periapical tissues (*DRB4*01:03*, *β* = 0.025, *P* = 7.4 × 10⁻⁶). Additionally, we discovered association signals at *DQA1*01:02* in diseases of lip and oral mucosa (*β* = –0.060, *P* = 5.4 × 10⁻⁵), and for *DQB1*03:01* in chronic apical periodontitis (*β* = 0.040, *P* = 6.3 × 10⁻⁶) (Supplementary Table 5). These results suggest that multiple oral phenotypes share common HLA-mediated inflammatory mechanisms.

### Shared heritability among oral traits

To understand the possible between-trait correlation, we computed genetic correlations using linkage disequilibrium score regression (LDSC) [19]. Of these, 378 pairs (17,1 %) showed statistically significant correlation (*P* < 0.05). As expected, strong positive correlations were observed between clinically and etiologically closely related phenotypes, such as caries and pulp and periapical diseases (*r_g_* = 0.84, 95% CI [0.78, 0.89], *P* = 5.0 × 10^-174^), pulpitis and chronic apical periodontitis (*r_g_* = 0.91, CI [0.82, 0.99], *P* = 1.3 × 10^-103^), tooth loss and chronic apical periodontitis (*r_g_* = 0.91, CI [0.76, 1.05], *P* = 1.7 × 10^-34^), and attrition and bruxism (*r_g_* = 0.90, CI [0.75, 1.04], *P* = 1.7 × 10^-33^). Strong positive correlations were also found between oral lichen planus and oral leukoplakia and other epithelial disorders (*r_g_* = 0.86, CI [0.50, 1.23], *P* = 3.3 × 10^-6^), and pulp necrosis and stomatitis (*r_g_* = 0.90, CI [0.35, 1.45], *P* = 0.0013).

Conversely, a small number of negative correlations were observed for occlusal phenotypes, such as cross bite and deep bite (*r_g_* = −0.51, CI [−0.78, −0.25], *P* = 0.0002), and cross bite and disto-occlusion (*r_g_* = −0.66, CI [-1.06, −0.25], *P* = 0.0014), (Supplementary Table 6).

Hierarchical clustering of genetic correlation profiles revealed distinct clusters of oral phenotypes that reflect both anatomical and etiological proximity. One inflammatory cluster included caries, apical periodontitis and periodontitis; a mucosal cluster grouped oral leukoplakia and lichen planus; and a structural/dentofacial cluster contained malocclusions and palatinal maxillary canine. These clusters are visualized in Fig. 4 and suggest substantial pleiotropy within oral tissues.

**Figure 4.**
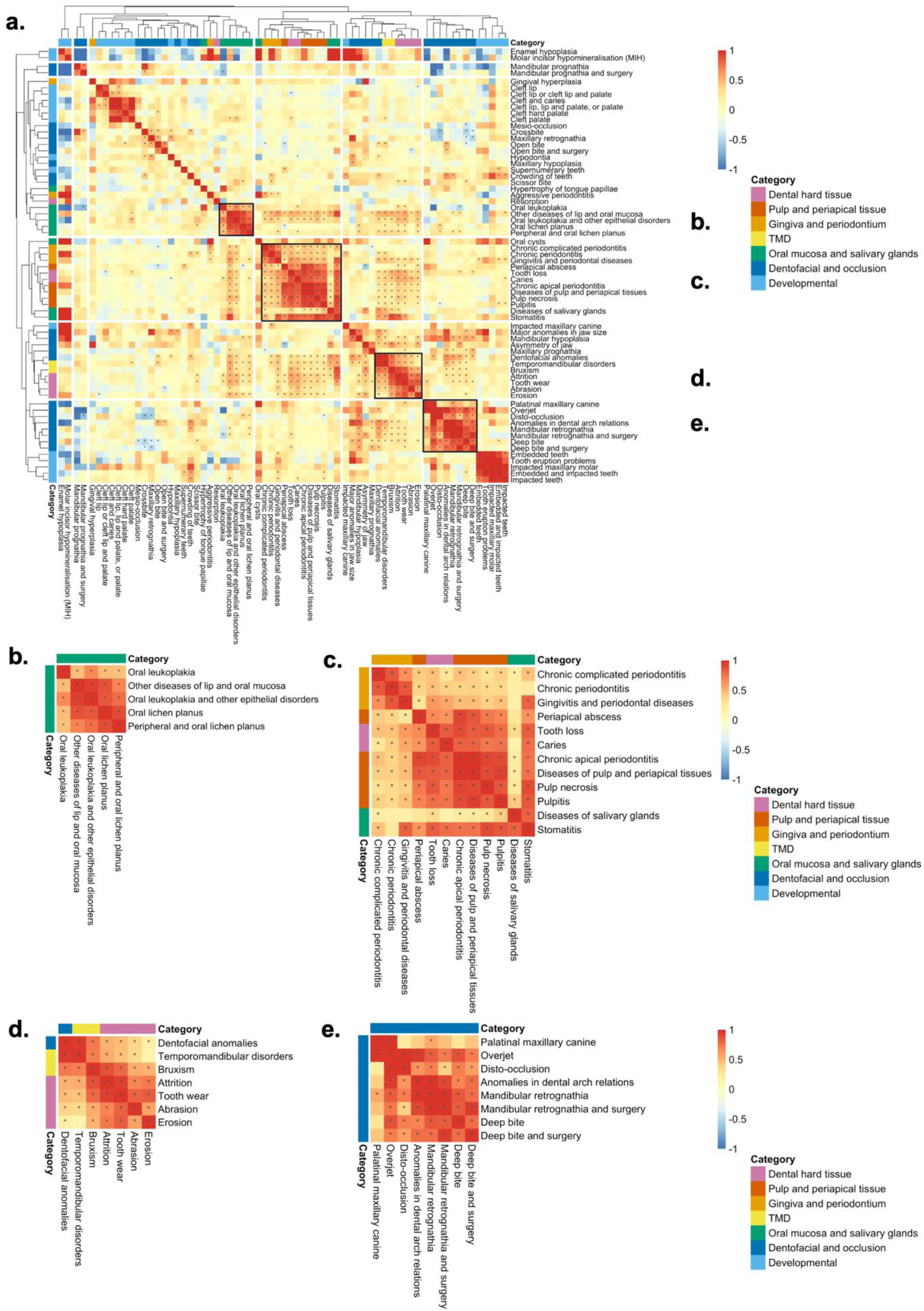
Heatmap of genetic correlations (*r_g_*) between oral phenotypes. a) Heatmap with genetic correlations between all oral phenotypes. Correlation values are clustered by hierarchical clustering with complete-linkage method, shown by dendrograms on both sides of the heatmap. The figure is annotated with phenotype categories. Statistically significant (P < 0.05) correlation values are marked with an asterisk (*). b-e) Zoomed-in views of clusters.

### Oral and systemic diseases share genetic architecture

To examine links between oral and overall health and disease burden, we calculated genetic correlations with 27 common systemic diseases and phenotypes using LDSC. Statistically significant (*P* < 0.05) positive genetic correlations were observed between 419 (25.4 %) pairs of oral phenotypes and systemic diseases, suggesting overlapping biological mechanisms that extend beyond the oral tissues (Supplementary Table 7).

The strongest genetic correlations were observed between chronic laryngitis/laryngotracheitis and gingivitis and periodontal diseases (*r_g_* = 0.97, 95% CI [0.58 – 1.36], *P* = 1.2 × 10^-6^), and caries (*r_g_* = 0.94, CI [0.58 – 1.30], *P* = 2.3 × 10^-7^). Several other inflammatory and infectious upper respiratory diseases were also strongly correlated with common oral diseases, forming a cluster in Fig. 5. For example, chronic rhinitis, nasopharyngitis and pharyngitis showed correlation with TMD (*r_g_* = 0.74, CI [0.60 – 0.88], *P* = 1.3 × 10^-24^) and diseases of pulp and periapical tissues (*r_g_* = 0.37, CI [0.27 – 0.48], *P* = 1.8 × 10^-12^). Additionally, genetic correlations were observed between autoimmune and oral diseases, such as seropositive rheumatoid arthritis and caries (*r_g_* = 0.24, CI [0.16 – 0.31], *P* = 9.4 × 10^-10^).

**Figure 5.**
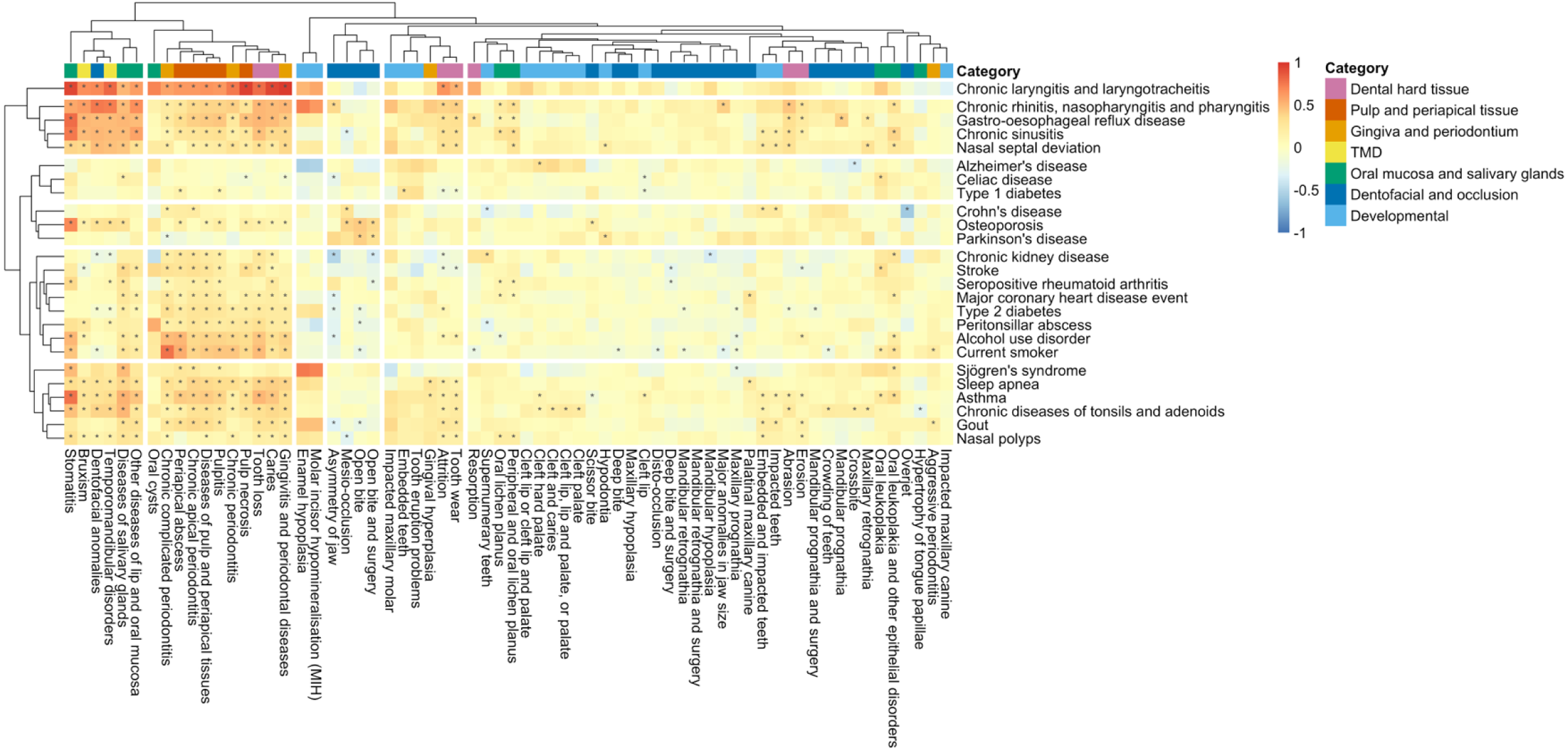
Heatmap of genetic correlations (*r_g_*) between all oral phenotypes and 27 systemic diseases and phenotypes. Correlation values are clustered by hierarchical clustering with complete-linkage method, shown by dendrograms on both sides of the heatmap. The figure is annotated with oral phenotype categories. Statistically significant (P < 0.05) correlation values are marked with an asterisk (*).

Chronic complicated periodontitis was genetically correlated with being a current smoker (*r_g_* = 0.77, CI [0.70 – 0.84], *P* = 1.1 × 10^-98^) and type 2 diabetes (*r_g_* = 0.33, CI [0.26 – 0.39], *P* = 1.1 × 10^-24^), aligning with previous epidemiological evidence linking periodontal inflammation to metabolic dysfunction and smoking [4, 20]. Positive correlation was also observed between bruxism and gastro-oesophageal reflux disease (*r_g_* = 0.51, CI [0.38 – 0.64], *P* = 1.1 × 10^-13^), as described in previous studies (Supplementary Table 7) [10].

### UK Biobank and Estonian Biobank data support key findings

To validate our GWAS results, we performed replication analyses with 12 oral phenotypes in UK Biobank summary statistics [21] and three oral phenotypes in Estonian Biobank data (Supplementary Tables 9 and 11). Replication in Estonian Biobank data was examined by conducting GWAS analyses for the three most common oral phenotypes, caries, gingivitis and periodontal diseases, and diseases of pulp and periapical tissues. The replication of lead and coding variants in the corresponding UK Biobank and Estonian Biobank phenotypes were tested. UK Biobank replication was observed for lead variants in hypodontia (rs2057579), chronic complicated periodontitis (rs2877160), and pulpitis (rs1292071) (Supplementary Table 10). In the Estonian biobank data, coding variants for caries (rs200486134 in *USP31*), gingivitis and periodontal diseases (rs16969968 in *CHRNA5,* rs1051730 *CHRNA3*), and pulp necrosis (rs2074204 in *MTMR3*) were replicated. Additionally, five lead variants were replicated in the summary statistics for diseases of pulp and periapical tissue, with similar effect sizes (Supplementary Table 12).

## Discussion

In this comprehensive genome-wide analysis of 67 oral and craniofacial phenotypes in over 500,000 individuals from the FinnGen study, we identified 105 genome-wide significant loci, of which 47 were not previously described in the literature in these oral and craniofacial phenotypes. The phenotypic spectrum analyzed spans dental hard tissues, oral mucosa, periodontal tissues, salivary glands and craniofacial structures, offering broader coverage than previous studies, which have largely focused on caries and periodontitis in smaller cohorts [5, 6]. By combining fine-mapping, HLA imputation and genetic correlation analyses, our work expands the known genetic architecture of oral diseases and their overlap with systemic conditions.

### Coding variants implicate key biological pathways

Among the 105 fine-mapped loci, we identified 14 coding variants, including six previously unreported variants. We identify shared pathways across dental phenotypes including ubiquitination, antigen presentation, lysosomal glycan catabolism, amino acid metabolism, epithelial integrity and bone formation. For example, a missense variant in *USP31* associated with caries (rs200486134), encodes a deubiquitinating enzyme linked to NF-κB activation and immune regulation [22]. Other members of the ubiquitin-specific protease family have also been implicated in dental biology; for example, *USP34* has been shown to influence odontogenic differentiation and tooth root morphogenesis through stabilization of *NFIC*, a transcription factor essential for root development [23]. Variant (rs75826658) in *MANBA*, associated with oral leukoplakia and epithelial disorders, implicate lysosomal β-mannosidase in glycan degradation and epithelial biology [24]. *THNSL2* (rs4129190), associated with hypodontia, encodes threonine synthase-like 2. An alternatively spliced isoform (SOFAT) of *THNSL2* has been identified in human T cells and shown to stimulate *IL-6* production and osteoclastogenesis [25]. *GPNMB* (rs11537976) linked here to TMD encodes Osteoactivin, a transmembrane glycoprotein. It promotes osteoblast differentiation and bone formation in vitro and in vivo (mouse models) and functions as an immunomodulator [26, 27]. Overall, these findings demonstrate that classical biological processes and particularly those in the ubiquitination, lysosomal glycan catabolism, amino acid metabolism, epithelial integrity, immunity and bone formation identified earlier from model organism and human disorder studies also contribute to common variation in dental and oral phenotypes.

### Finnish-enriched variants and insights from a founder population

Several variants were enriched in the Finnish population, reflecting its unique genetic architecture shaped by historical bottlenecks and drift [14, 28]. These included variants near *HDGFL1* (encoding a growth factor–like protein implicated in cell proliferation), *LIPA* (encoding lysosomal acid lipase involved in lipid metabolism and inflammatory processes), *NUP42* (encoding a nucleoporin required for mRNA export), and *MBP* (encoding myelin basic protein essential for myelination), associated respectively with peripheral/oral lichen planus, pulpitis, TMD, and anomalies of dental arch relations. Such population-enriched variants, some absent in non-Finnish Europeans, may implicate biological pathways not previously linked to oral and craniofacial phenotypes and highlight the value of founder populations in rare variant discovery.

### HLA region associations with oral mucosal disease

Our HLA fine-mapping identified strong and specific associations between class I and class II alleles and diverse oral phenotypes. The strongest signals were for oral and peripheral lichen planus, with *DQB1*05:01* and *DQA1*01:01* showing large positive associations, and *DQA1*01:02* showing a protective association with diseases of the lip and oral mucosa as described earlier [11]. Inflammatory dental phenotypes, including pulpitis and chronic apical periodontitis, were associated with *DRB4*01:03* and *DQB1*03:01*, respectively, and attrition was associated with *B*18:01*. These findings are consistent with previous studies linking HLA variation to lichen planus and pulpal diseases suggesting that antigen presentation may influence susceptibility to a broad range of oral diseases [11, 13].

### Genetic correlations among oral traits and with systemic conditions

Our genome-wide genetic correlation analyses revealed extensive overlap both within oral phenotypes and between oral and systemic diseases. As expected, oral phenotypes that are clinically and etiologically linked showed the strongest positive correlations, such as caries with pulp and periapical diseases, pulpitis with chronic apical periodontitis, and attrition with bruxism. Mucosal disorders, including oral lichen planus and oral leukoplakia, also displayed strong genetic overlap, consistent with previous evidence that both conditions involve epithelial and immune dysregulation [29]. Conversely, some malocclusion traits exhibited negative correlations. For example, cross bite with deep bite reflecting contrasting dentofacial patterns that are less commonly observed together. Hierarchical clustering of correlation profiles delineated three main oral disease clusters: an inflammatory dental cluster (e.g., caries, apical periodontitis, periodontitis), a mucosal cluster (e.g., oral leukoplakia, lichen planus), and a structural/dentofacial cluster (e.g., malocclusions).

Extending the analysis to 27 common systemic diseases and traits revealed that nearly one-quarter of oral–systemic pairs were genetically correlated. Strongest overlaps were seen between chronic laryngitis/laryngotracheitis and both gingivitis/periodontal disease and caries, as well as between upper respiratory infections and TMD or diseases of pulp and periapical tissues. Autoimmune conditions such as seropositive rheumatoid arthritis showed positive correlations with caries, in line with prior evidence linking systemic immune dysregulation to oral pathology [4]. Chronic complicated periodontitis also correlated strongly with current smoking and type 2 diabetes, reinforcing well-established epidemiological links between periodontal inflammation, smoking, and metabolic dysfunction [30, 31]. Furthermore, bruxism was genetically correlated with gastro-oesophageal reflux disease, echoing previous clinical observations [32].

### Limitations

This study has several limitations. Using phenotype data derived from registries may lead to misclassification or under-detection of certain diagnoses, particularly rare or subclinical conditions. Although the Finnish founder population enhances power to detect low-frequency variants, generalizability of population-specific signals may be limited, and replication in non-Finnish cohorts is more challenging. Finally, while our analyses suggest mechanistic hypotheses, functional validation of implicated variants is needed to establish causal relationships.

## Methods

### Study cohort

FinnGen is a nationwide research project that integrates genetic and health registry data from over 500,000 individuals in Finland. It combines extensive genetic data with comprehensive longitudinal health data, enabling large-scale GWAS on a wide range of diseases, including oral and systemic conditions. [14]

We utilized data from FinnGen Release R12, which includes genetic and health registry data from over 500,000 individuals (N = 500 348). Oral disease phenotypes were identified based on both ICD-10 and ICD-9 codes, covering 67 conditions, including diseases of the dental hard tissues, periodontium, oral mucosa, salivary glands, jaw structure, occlusion, and other related oral and craniofacial disorders. Detailed phenotype definitions and case-control numbers are provided in Supplementary Table 1.

The number of individuals per phenotype category (Fig. 1b) was calculated by identifying all unique individuals in the case cohorts of phenotypes in the categories. Individuals were included only once within each phenotype category. Similarly, the number of fine-mapped GWAS hits per category (Fig. 1c) was determined by calculating the number of unique loci within each phenotype category.

### Genotyping and sample quality control and imputation

Genotyping in the FinnGen cohort was performed by using Illumina (Illumina Inc., San Diego, CA, USA) and Affymetrix arrays (Thermo Fisher Scientific, Santa Clara, CA, USA) and lifted over to Genome Reference Consortium Human Build version 38 (GRCh38/hg38). Individuals with high genotype absence (> 5 %), inexplicit sex or excess heterozygosity (± 4 standard deviations) were excluded from the data [14]. Additionally, variants that had high absence (> 2 %), low minor allele count (< 3) or low Hardy-Weinberg Equilibrium (HWE) (*P* < 1 × 10^−6^) were removed. All individuals in the cohort were Finns and matched against the SiSu v4 reference panel (http://www.sisuproject.fi/).

Before imputation, array-genotyped samples were pre-phased with Eagle 2.3.5 using the default parameters, except for the number of conditioning haplotypes, which was set to 20 000.

Genotype imputation was carried out by using the population-specific SISu v4.2 imputation reference panel with Beagle 4.1 (version 27Jan18.7e1). Post-imputation quality control involved checking the expected conformity of the imputation INFO-value distribution, minor allele frequency (MAF) differences between the target dataset and the imputation reference panel and checking chromosomal continuity of the imputed genotype calls.

### Genome-wide association analysis and fine-mapping

Genome-wide association testing was conducted using the Regenie v2.2.4 software and the FinnGen Regenie pipeline (https://github.com/FINNGEN/regenie-pipelines). The analysis was adjusted for current age or age at death, sex, genotyping chip, genetic relationship, and the first 10 principal components. Genome-wide significant associations were defined using a statistical significance threshold of *P* < 5 × 10⁻⁸. To explore the causality of genomic variations in the loci related to the corresponding phenotype, a fine-mapping approach was employed using the SuSiE “Sum of Single Effects” model [16]. The variants were annotated with the nearest protein coding gene based on the Ensembl database (release 114) [33].

### HLA fine-mapping

HLA fine-mapping was conducted using multivariate logistic regression models with covariates including age, sex, cohort, and the first 10 genetic principal components (PCs). Imputed HLA allele dosages were obtained from the FinnGen reference panel, and rare alleles with MAF < 1% were excluded from the analysis.

### Cross-trait genetic correlation analysis

We performed cross-trait genetic correlation analyses using linkage disequilibrium score regression (LDSC). Summary statistics were derived from FinnGen GWAS results for each oral phenotype. SNP-based heritability estimates are presented on the observed scale, based on the European LD score reference panel [19] (Supplementary Table 8). Heatmaps (Fig. 4-5) were created using pheatmap -package in R. Complete-linkage method was used for hierarchical clustering.

### Definition of previously unreported loci and variants using GWAS Catalog cross-referencing

New GWAS loci were defined according to fine-mapping loci regions. NHGRI-EBI GWAS Catalog variant associations were downloaded (v1.0, 2025-09-15), and all variants located within the loci regions were searched and filtered for previous genome-wide significant associations with oral or craniofacial phenotypes in the same phenotype category. Diseases of dental hard tissue and pulp and periapical tissue were combined due to etiological similarities. Lead variants and credible set coding variants were searched from GWAS Catalog to determine any previous GWAS associations. [17]

### Replication

Replication of lead and coding variants of FinnGen oral phenotypes was performed using summary statistics from 12 corresponding UK Biobank traits, obtained from previously published GWAS studies listed in Supplementary Table 9. In addition, three GWAS analyses were conducted in the Estonian Biobank for replication (Supplementary Table 11). The replication results are summarized in Supplementary Tables 10 and 12. A statistical significance threshold of *P* < 0.05 was applied to define successful replication.

## Supporting information

FinnGen_banner

## Ethics statement

Study subjects in FinnGen provided informed consent for biobank research, based on the Finnish Biobank Act. Alternatively, separate research cohorts, collected prior the Finnish Biobank Act came into effect (in September 2013) and start of FinnGen (August 2017), were collected based on study-specific consents and later transferred to the Finnish biobanks after approval by Fimea (Finnish Medicines Agency), the National Supervisory Authority for Welfare and Health. Recruitment protocols followed the biobank protocols approved by Fimea. The Coordinating Ethics Committee of the Hospital District of Helsinki and Uusimaa (HUS) statement number for the FinnGen study is Nr HUS/990/2017.

The FinnGen study is approved by Finnish Institute for Health and Welfare (permit numbers: THL/2031/6.02.00/2017, THL/1101/5.05.00/2017, THL/341/6.02.00/2018, THL/2222/6.02.00/2018, THL/283/6.02.00/2019, THL/1721/5.05.00/2019 and THL/1524/5.05.00/2020), Digital and population data service agency (permit numbers: VRK43431/2017-3, VRK/6909/2018-3, VRK/4415/2019-3), the Social Insurance Institution (permit numbers: KELA 58/522/2017, KELA 131/522/2018, KELA 70/522/2019, KELA 98/522/2019, KELA 134/522/2019, KELA 138/522/2019, KELA 2/522/2020, KELA 16/522/2020), Findata permit numbers THL/2364/14.02/2020, THL/4055/14.06.00/2020, THL/3433/14.06.00/2020, THL/4432/14.06/2020, THL/5189/14.06/2020, THL/5894/14.06.00/2020, THL/6619/14.06.00/2020, THL/209/14.06.00/2021, THL/688/14.06.00/2021, THL/1284/14.06.00/2021, THL/1965/14.06.00/2021, THL/5546/14.02.00/2020, THL/2658/14.06.00/2021, THL/4235/14.06.00/2021, THL/4990/14.02.00/2023 Statistics Finland (permit numbers: TK-53-1041-17 and TK/143/07.03.00/2020 (earlier TK-53-90-20) TK/1735/07.03.00/2021, TK/3112/07.03.00/2021) and Finnish Registry for Kidney Diseases permission/extract from the meeting minutes on 4th July 2019.

The Biobank Access Decisions for FinnGen samples and data utilized in FinnGen Data Freeze 13 include: THL Biobank BB2017_55, BB2017_111, BB2018_19, BB_2018_34, BB_2018_67, BB2018_71, BB2019_7, BB2019_8, BB2019_26, BB2020_1, BB2021_65, BB22-0025-A01, BB22-0025-A03, BB23-0222-A01, BB22-0025-A04, BB22-0025-A06, BB22-0025-A08, THLBB2024_30. Finnish Red Cross Blood Service Biobank 7.12.2017, 13.11.2023, 001-2023, Helsinki Biobank HUS/359/2017, HUS/248/2020, HUS/430/2021 §28, §29, HUS/150/2022 §12, §13, §14, §15, §16, §17, §18, §23, §58, §59, HUS/128/2023 §18, BB22-0025-A01, BB22-0025-A02, BB22-0025-A05, BB22-0025-A07, BB22-0025-A09, BB22-0025-A10, BB22-0025-A03, BB23-0222-A01, BB22-0025-A04, BB22-0025-A06, BB22-0025-A08, Amendment_BB22-0025-A05, Decision allowing to continue data processing until 31st Aug 2027: BB_2021-0140, HUS/150/2022 §12, BB_2021-0139, HUS/150/2022 §13, BB_2021-0161,HUS/150/2022 §14, BB_2021-0164, HUS/150/2022 §15, BB_2021-0169, HUS/150/2022 §16, BB_2021-0170, HUS/150/2022 §17, BB_2021-0179, HUS/150/2022 §18, BB_2022-0262, HUS/150/2022 §58, BB22-0067, HUS/150/2022 §59, Auria Biobank AB17-5154 and amendment #1 (August 17 2020) and amendments BB_2021-0140, BB_2021-0156 (August 26 2021, Feb 2 2022), BB_2021-0169, BB_2021-0179, BB_2021-0161, AB20-5926 and amendment #1 (April 23 2020) and it’s modifications (Sep 22 2021), BB_2022-0262, BB_2022-0256, BB22-0025-A01, BB22-0025-A02, BB22-0025-A03, BB23-0222_A01, BB22-0025-A02, BB22-0025-A05, BB22-0025-A07, BB22-0025-A09, BB22-0025-A10, BB22-0025-A03, BB23-0222-A01, BB22-0025-A04, BB22-0025-A06, BB22-0025-A08, Decision allowing to continue data processing until 31st Aug 2027: AB20-5926, BB_2021-0140, BB_2021-0156, BB_2021-0161, BB_2021-0161, BB_2021-0164, BB_2021-0169, BB_2021-0179, BB_2022-0262, Biobank Borealis of Northern Finland_2017_1013, 2021_5010, 2021_5010 Amendment, 2021_5018, 2021_5018 Amendment, 2021_5015, 2021_5015 Amendment, 2021_5015 Amendment_2, 2021_5023, 2021_5023 Amendment, 2021_5023 Amendment_2, 2021_5017, 2021_5017 Amendment, 2022_6001, 2022_6001 Amendment, 2022_6006 Amendment, 2022_6006 Amendment_2, BB22-0067, 2022_0262, 2022_0262 Amendment, BB22-0025-A01, BB22-0025-A02, BB22-0025-A05, BB22-0025-A07, BB22-0025-A09, BB22-0025-A10, BB22-0025-A03, BB23-0222-A01, BB22-0025-A04, BB22-0025-A06, BB22-0025-A08, Decision allowing to continue data processing until 31st Aug 2027: BB/2021/5015, BB/2021/5017, BB/2021/5018, BB/2021/5023, BB/2022/6006, BB/2022/6001, BB/2022-0262, BB/2021/5010, Biobank of Eastern Finland 1186/2018 and amendment 22§/2020, 53§/2021, 13§/2022, 14§/2022, 15§/2022, 27§/2022, 28§/2022, 29§/2022, 33§/2022, 35§/2022, 36§/2022, 37§/2022, 39§/2022, 7§/2023, 32§/2023, 33§/2023, 34§/2023, 35§/2023, 36§/2023, 37§/2023, 38§/2023, 39§/2023, 40§/2023, 41§/2023, BB22-0025-A01, BB22-0025-A02, BB22-0025-A05, BB22-0025-A07, BB22-0025-A09, BB22-0025-A10, BB22-0025-A03, BB23-0222-A01, BB22-0025-A04, BB22-0025-A06, BB22-0025-A08, Decision allowing to continue data processing until 31st Aug 2027: MO-BB_2021-0179-A0, MO-BB_2021-0156_PRE-A01, BB_2021-0140, MO-BB_2021-0170_PRE-A0, MO-BB_2021-0169-A01, MO-BB_2022-0256-A01, MO-BB_2021-0161-A01, MO-BB_2021-0161-A02, BB22-0067-A01, MO-BB_2022-0262-A0, Finnish Clinical Biobank Tampere MH0004 and amendments (21.02.2020 & 06.10.2020), BB2021-0140 8§/2021, 9§/2021, §9/2022, §10/2022, §12/2022, 13§/2022, §20/2022, §21/2022, §22/2022, §23/2022, 28§/2022, 29§/2022, 30§/2022, 31§/2022, 32§/2022, 38§/2022, 40§/2022, 42§/2022, 1§/2023, BB2021-0140, BB22-0025-A01, BB_2021-0161, BB22-0025-A02, BB22-0025-A05, BB22-0025-A07, BB22-0025-A09, BB22-0025-A10, BB22-0025-A03, BB23-0222-A01, BB22-0025-A04, BB22-0025-A06, BB22-0025-A08, Decision allowing to continue data processing until 31st Aug 2027: BB_2021-0140, BB_ 2021-0161, BB_ 2021-0179, BB_ 2021-0156, BB_ 2021-0169, BB_ 2021-0170, BB22-0067-A01, Central Finland Biobank 1-2017, BB_2021-0169, BB_2021-0179, BB_2022-0256, BB_2022-0262, Decision allowing to continue data processing until 31st Aug 2027 for projects: BB_2021-0179, BB22-0067,BB_2022-0262, BB_2021-0170, BB_2021-0164, BB_2021-0161, and BB_2021-0169, BB22-0025-A01, BB22-0025-A02, BB22-0025-A05, BB22-0025-A07, BB22-0025-A09, BB22-0025-A10, BB22-0025-A03, BB23-0222-A01, BB22-0025-A04, BB22-0025-A06, BB22-0025-A08, Terveystalo Biobank STB 2018001 and amendment 25th Aug 2020, Finnish Hematological Registry and Clinical Biobank decision 18th June 2021, Amendment 2nd January 2024 and Arctic biobank P0844: ARC_2021_1001, ARC_2023_3003 (BB22-0025-A01), BB22-0025-A03, BB23-0222-A01, BB22-0025-A04, BB22-0025-A06, BB22-0025-A08.

The Estonian Biobank is a volunteer-based biobank with 212,955 participants in the current data freeze [PMID:40188112]. All biobank participants have signed a broad informed consent form and information on ICD-10 codes is obtained via regular linking with the national Health Insurance Fund and other relevant databases, with majority of the electronic health records having been collected since 2004 [PMID:24518929]. Analyses were restricted to individuals with European ancestry. The activities of the EstBB are regulated by the Human Genes Research Act, which was adopted in 2000 specifically for the operations of EstBB. Individual level data analysis in EstBB was carried out under ethical approval 1.1-12/624 from the Estonian Committee on Bioethics and Human Research (Estonian Ministry of Social Affairs), using data according to release application 6-7/GI/2014 from the Estonian Biobank.

This study was conducted using publicly available genome-wide association summary statistics derived from the UK Biobank resource under approved data access. All UK Biobank participants provided written informed consent for their data to be used in health-related research. The UK Biobank has obtained ethical approval from the North West Multi-centre Research Ethics Committee (REC reference: 21/NW/0157).

## Data Availability

All data produced in the present study are available upon reasonable request to the authors.

## Acknowledgements

We want to acknowledge the participants and investigators of the FinnGen study. The FinnGen project is funded by two grants from Business Finland (HUS 4685/31/2016 and UH 4386/31/2016) and the following industry partners: AbbVie Inc., AstraZeneca UK Ltd, Biogen MA Inc., Bristol Myers Squibb Inc. (and Celgene Corporation & Celgene International II Sàrl), Genentech Inc., Merck Sharp & Dohme LCC, Pfizer Inc., GlaxoSmithKline Intellectual Property Development Ltd., Sanofi US Services Inc., Maze Therapeutics Inc., Johnson&Johnson Innovative Medicine Inc., Novartis AG, Boehringer Ingelheim International GmbH and Bayer AG. Following biobanks are acknowledged for delivering biobank samples to FinnGen: Auria Biobank (www.auria.fi/biopankki), THL Biobank (www.thl.fi/biobank), Helsinki Biobank (www.helsinginbiopankki.fi), Biobank Borealis of Northern Finland (https://www.ppshp.fi/Tutkimus-ja-opetus/Biopankki/Pages/Biobank-Borealis-briefly-in-English.aspx), Finnish Clinical Biobank Tampere (www.tays.fi/en-US/Research_and_development/Finnish_Clinical_Biobank_Tampere), Biobank of Eastern Finland (www.ita-suomenbiopankki.fi/en), Central Finland Biobank (www.ksshp.fi/fi-FI/Potilaalle/Biopankki), Finnish Red Cross Blood Service Biobank (www.veripalvelu.fi/verenluovutus/biopankkitoiminta), Terveystalo Biobank (www.terveystalo.com/fi/Yritystietoa/Terveystalo-Biopankki/Biopankki/) and Arctic Biobank (https://www.oulu.fi/en/university/faculties-and-units/faculty-medicine/northern-finland-birth-cohorts-and-arctic-biobank). All Finnish Biobanks are members of BBMRI.fi infrastructure (https://www.bbmri-eric.eu/national-nodes/finland/). Finnish Biobank Cooperative -FINBB (https://finbb.fi/) is the coordinator of BBMRI-ERIC operations in Finland. The Finnish biobank data can be accessed through the Fingenious® services (https://site.fingenious.fi/en/) managed by FINBB. The Estonian Genome Center analyses were partially carried out in the High Performance Computing Center, University of Tartu. The Estonian Biobank Research Team was responsible for data collection, genotyping, quality control and imputation and consisted of Andres Metspalu, Mait Metspalu, Lili Milani, Reedik Mägi, Mari Nelis, Tõnu Esko and Georgi Hudjashov.

## Funding

Supported by Finska Läkaresällskapet (to KK and NM); and Academy of Finland (grant number 355567 to NM). The work of the Estonian Genome Center, University of Tartu was funded by the European Union through Horizon Europe research and innovation programs under grants no. 101137201 and 101137154, and Estonian Research Council Grant PRG1291.

## Notes

### Competing Interest Statement

The authors have declared no competing interest.

### Funding Statement

Supported by Finska Laekaresaellskapet (to KK and NM); and Academy of Finland (grant number 355567 to NM). The work of the Estonian Genome Center, University of Tartu was funded by the European Union through Horizon Europe research and innovation programs under grants no. 101137201 and 101137154, and Estonian Research Council Grant PRG1291.

### Author Declarations

Study subjects in FinnGen provided informed consent for biobank research, based on the Finnish Biobank Act. Alternatively, separate research cohorts, collected prior the Finnish Biobank Act came into effect (in September 2013) and start of FinnGen (August 2017), were collected based on study-specific consents and later transferred to the Finnish biobanks after approval by Fimea (Finnish Medicines Agency), the National Supervisory Authority for Welfare and Health. Recruitment protocols followed the biobank protocols approved by Fimea. The Coordinating Ethics Committee of the Hospital District of Helsinki and Uusimaa (HUS) statement number for the FinnGen study is Nr HUS/990/2017. The FinnGen study is approved by Finnish Institute for Health and Welfare (permit numbers: THL/2031/6.02.00/2017, THL/1101/5.05.00/2017, THL/341/6.02.00/2018, THL/2222/6.02.00/2018, THL/283/6.02.00/2019, THL/1721/5.05.00/2019 and THL/1524/5.05.00/2020), Digital and population data service agency (permit numbers: VRK43431/2017-3, VRK/6909/2018-3, VRK/4415/2019-3), the Social Insurance Institution (permit numbers: KELA 58/522/2017, KELA 131/522/2018, KELA 70/522/2019, KELA 98/522/2019, KELA 134/522/2019, KELA 138/522/2019, KELA 2/522/2020, KELA 16/522/2020), Findata permit numbers THL/2364/14.02/2020, THL/4055/14.06.00/2020, THL/3433/14.06.00/2020, THL/4432/14.06/2020, THL/5189/14.06/2020, THL/5894/14.06.00/2020, THL/6619/14.06.00/2020, THL/209/14.06.00/2021, THL/688/14.06.00/2021, THL/1284/14.06.00/2021, THL/1965/14.06.00/2021, THL/5546/14.02.00/2020, THL/2658/14.06.00/2021, THL/4235/14.06.00/2021, THL/4990/14.02.00/2023 Statistics Finland (permit numbers: TK-53-1041-17 and TK/143/07.03.00/2020 (earlier TK-53-90-20) TK/1735/07.03.00/2021, TK/3112/07.03.00/2021) and Finnish Registry for Kidney Diseases permission/extract from the meeting minutes on 4th July 2019. The Biobank Access Decisions for FinnGen samples and data utilized in FinnGen Data Freeze 13 include: THL Biobank BB2017_55, BB2017_111, BB2018_19, BB_2018_34, BB_2018_67, BB2018_71, BB2019_7, BB2019_8, BB2019_26, BB2020_1, BB2021_65, BB22-0025-A01, BB22-0025-A03, BB23-0222-A01, BB22-0025-A04, BB22-0025-A06, BB22-0025-A08, THLBB2024_30. Finnish Red Cross Blood Service Biobank 7.12.2017, 13.11.2023, 001-2023, Helsinki Biobank HUS/359/2017, HUS/248/2020, HUS/430/2021 28, 29, HUS/150/2022 12, 13, 14, 15, 16, 17, 18, 23, 58, 59, HUS/128/2023 18, BB22-0025-A01, BB22-0025-A02, BB22-0025-A05, BB22-0025-A07, BB22-0025-A09, BB22-0025-A10, BB22-0025-A03, BB23-0222-A01, BB22-0025-A04, BB22-0025-A06, BB22-0025-A08, Amendment_BB22-0025-A05, Decision allowing to continue data processing until 31st Aug 2027: BB_2021-0140, HUS/150/2022 12, BB_2021-0139, HUS/150/2022 13, BB_2021-0161, HUS/150/2022 14, BB_2021-0164, HUS/150/2022 15, BB_2021-0169, HUS/150/2022 16, BB_2021-0170, HUS/150/2022 17, BB_2021-0179, HUS/150/2022 18, BB_2022-0262, HUS/150/2022 58, BB22-0067, HUS/150/2022 59, Auria Biobank AB17-5154 and amendment #1 (August 17 2020) and amendments BB_2021-0140, BB_2021-0156 (August 26 2021, Feb 2 2022), BB_2021-0169, BB_2021-0179, BB_2021-0161, AB20-5926 and amendment #1 (April 23 2020) and its modifications (Sep 22 2021), BB_2022-0262, BB_2022-0256, BB22-0025-A01, BB22-0025-A02, BB22-0025-A03, BB23-0222_A01, BB22-0025-A02, BB22-0025-A05, BB22-0025-A07, BB22-0025-A09, BB22-0025-A10, BB22-0025-A03, BB23-0222-A01, BB22-0025-A04, BB22-0025-A06, BB22-0025-A08, Decision allowing to continue data processing until 31st Aug 2027: AB20-5926, BB_2021-0140, BB_2021-0156, BB_2021-0161, BB_2021-0161, BB_2021-0164, BB_2021-0169, BB_2021-0179, BB_2022-0262, Biobank Borealis of Northern Finland_2017_1013, 2021_5010, 2021_5010 Amendment, 2021_5018, 2021_5018 Amendment, 2021_5015, 2021_5015 Amendment, 2021_5015 Amendment_2, 2021_5023, 2021_5023 Amendment, 2021_5023 Amendment_2, 2021_5017, 2021_5017 Amendment, 2022_6001, 2022_6001 Amendment, 2022_6006 Amendment, 2022_6006 Amendment_2, BB22-0067, 2022_0262, 2022_0262 Amendment, BB22-0025-A01, BB22-0025-A02, BB22-0025-A05, BB22-0025-A07, BB22-0025-A09, BB22-0025-A10, BB22-0025-A03, BB23-0222-A01, BB22-0025-A04, BB22-0025-A06, BB22-0025-A08, Decision allowing to continue data processing until 31st Aug 2027: BB/2021/5015, BB/2021/5017, BB/2021/5018, BB/2021/5023, BB/2022/6006, BB/2022/6001, BB/2022-0262, BB/2021/5010, Biobank of Eastern Finland 1186/2018 and amendment 22/2020, 53/2021, 13/2022, 14/2022, 15/2022, 27/2022, 28/2022, 29/2022, 33/2022, 35/2022, 36/2022, 37/2022, 39/2022, 7/2023, 32/2023, 33/2023, 34/2023, 35/2023, 36/2023, 37/2023, 38/2023, 39/2023, 40/2023, 41/2023, BB22-0025-A01, BB22-0025-A02, BB22-0025-A05, BB22-0025-A07, BB22-0025-A09, BB22-0025-A10, BB22-0025-A03, BB23-0222-A01, BB22-0025-A04, BB22-0025-A06, BB22-0025-A08, Decision allowing to continue data processing until 31st Aug 2027: MO-BB_2021-0179-A0, MO-BB_2021-0156_PRE-A01, BB_2021-0140, MO-BB_2021-0170_PRE-A0, MO-BB_2021-0169-A01, MO-BB_2022-0256-A01, MO-BB_2021-0161-A01, MO-BB_2021-0161-A02, BB22-0067-A01, MO-BB_2022-0262-A0, Finnish Clinical Biobank Tampere MH0004 and amendments (21.02.2020 & 06.10.2020), BB2021-0140 8/2021, 9/2021, 9/2022, 10/2022, 12/2022, 13/2022, 20/2022, 21/2022, 22/2022, 23/2022, 28/2022, 29/2022, 30/2022, 31/2022, 32/2022, 38/2022, 40/2022, 42/2022, 1/2023, BB2021-0140, BB22-0025-A01, BB_2021-0161, BB22-0025-A02, BB22-0025-A05, BB22-0025-A07, BB22-0025-A09, BB22-0025-A10, BB22-0025-A03, BB23-0222-A01, BB22-0025-A04, BB22-0025-A06, BB22-0025-A08, Decision allowing to continue data processing until 31st Aug 2027: BB_2021-0140, BB_ 2021-0161, BB_ 2021-0179, BB_ 2021-0156, BB_ 2021-0169, BB_ 2021-0170, BB22-0067-A01, Central Finland Biobank 1-2017, BB_2021-0169, BB_2021-0179, BB_2022-0256, BB_2022-0262, Decision allowing to continue data processing until 31st Aug 2027 for projects: BB_2021-0179, BB22-0067,BB_2022-0262, BB_2021-0170, BB_2021-0164, BB_2021-0161, and BB_2021-0169, BB22-0025-A01, BB22-0025-A02, BB22-0025-A05, BB22-0025-A07, BB22-0025-A09, BB22-0025-A10, BB22-0025-A03, BB23-0222-A01, BB22-0025-A04, BB22-0025-A06, BB22-0025-A08, Terveystalo Biobank STB 2018001 and amendment 25th Aug 2020, Finnish Hematological Registry and Clinical Biobank decision 18th June 2021, Amendment 2nd January 2024 and Arctic biobank P0844: ARC_2021_1001, ARC_2023_3003 (BB22-0025-A01), BB22-0025-A03, BB23-0222-A01, BB22-0025-A04, BB22-0025-A06, BB22-0025-A08. The Estonian Biobank is a volunteer-based biobank with 212,955 participants in the current data freeze [PMID:40188112]. All biobank participants have signed a broad informed consent form and information on ICD-10 codes is obtained via regular linking with the national Health Insurance Fund and other relevant databases, with majority of the electronic health records having been collected since 2004 [PMID:24518929]. Analyses were restricted to individuals with European ancestry. The activities of the EstBB are regulated by the Human Genes Research Act, which was adopted in 2000 specifically for the operations of EstBB. Individual level data analysis in EstBB was carried out under ethical approval 1.1-12/624 from the Estonian Committee on Bioethics and Human Research (Estonian Ministry of Social Affairs), using data according to release application 6-7/GI/2014 from the Estonian Biobank. This study was conducted using publicly available genome-wide association summary statistics derived from the UK Biobank resource under approved data access. All UK Biobank participants provided written informed consent for their data to be used in health-related research. The UK Biobank has obtained ethical approval from the North West Multi-centre Research Ethics Committee (REC reference: 21/NW/0157).

