## Supplementary_Figures1 for "Genetic architecture of 67 oral diseases and their links to systemic diseases"

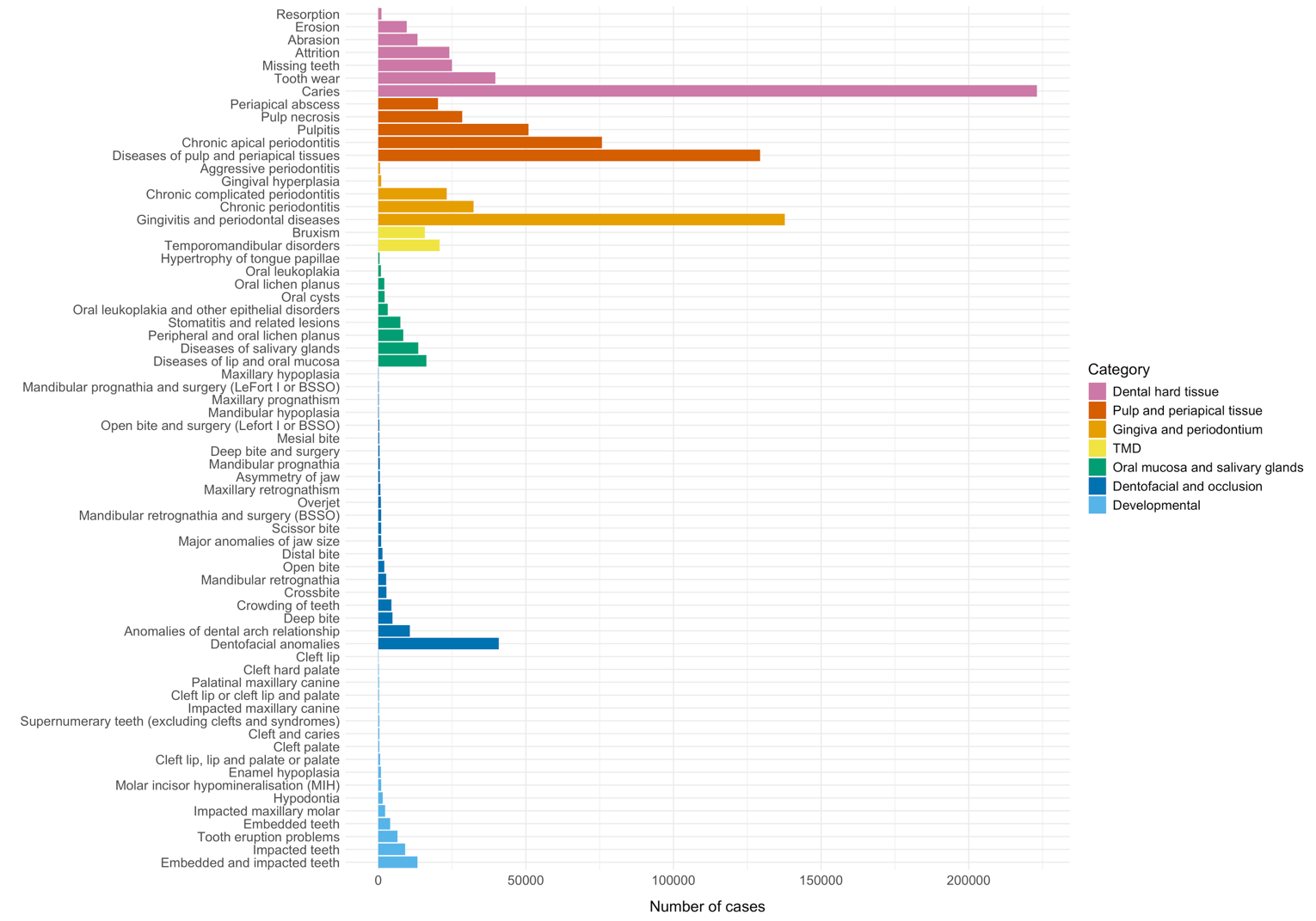
Figure S1. Number of cases of all oral phenotypes. *Phenotypes are organized according to phenotype categories.*

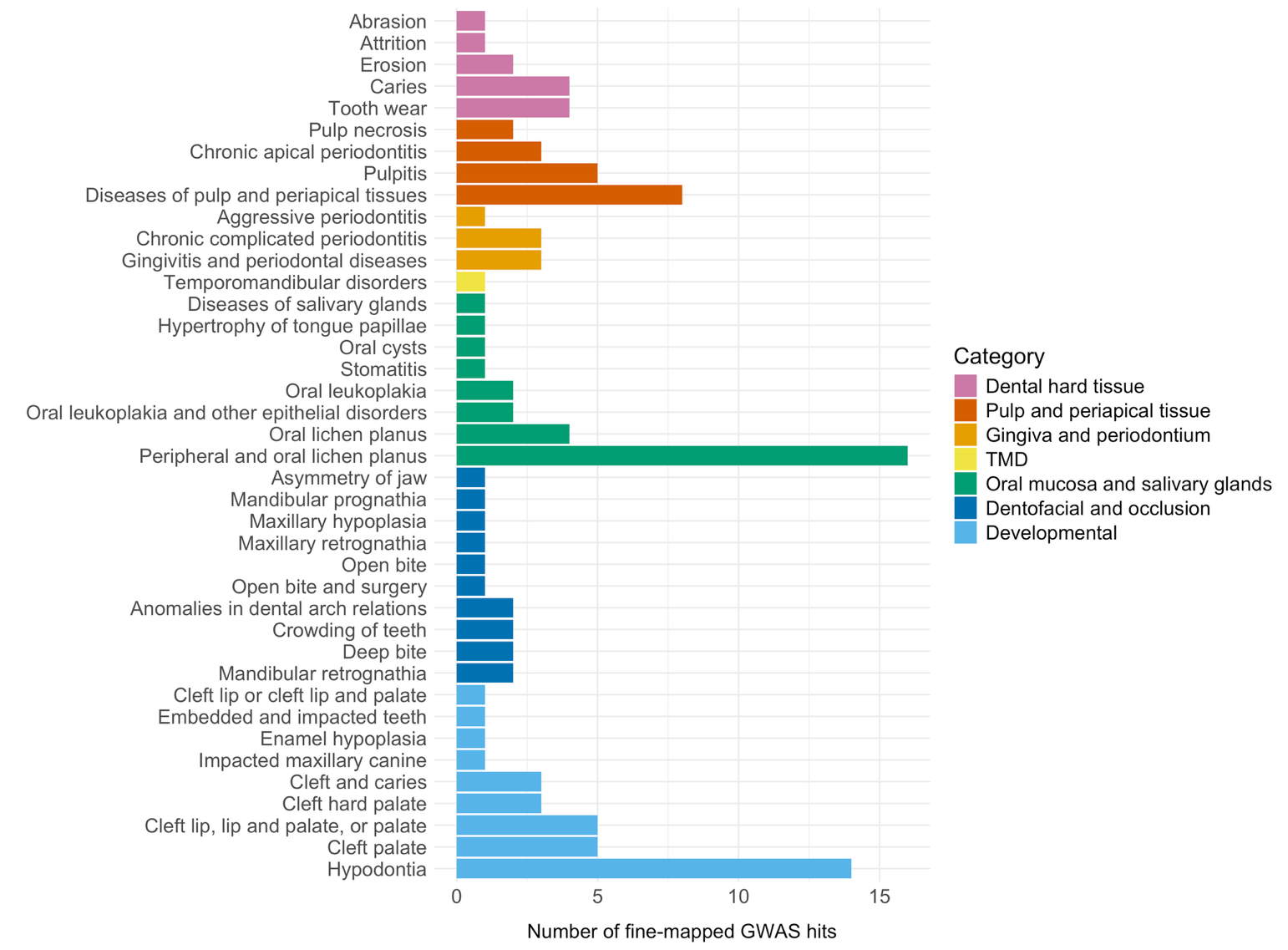

Figure S2. Number of fine-mapped GWAS hits per oral phenotype. *Phenotypes are organized according to phenotype categories.*
