## Supplementary_Figures2 for "Genetic architecture of 67 oral diseases and their links to systemic diseases"

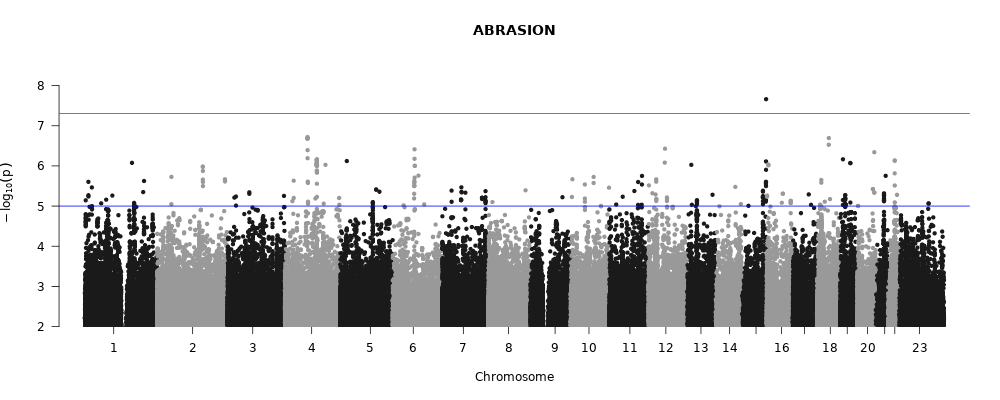

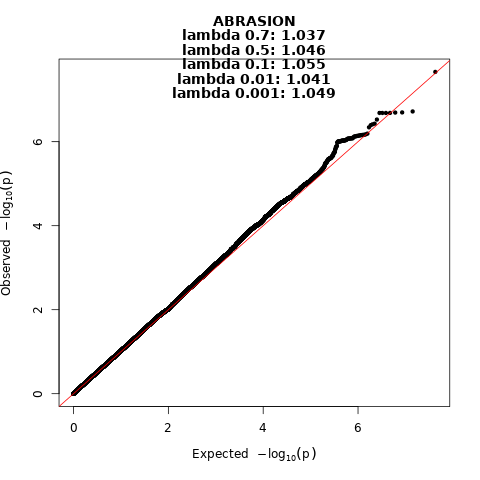

Figure 1. Manhattan and QQ-plot of phenotype Abrasion.

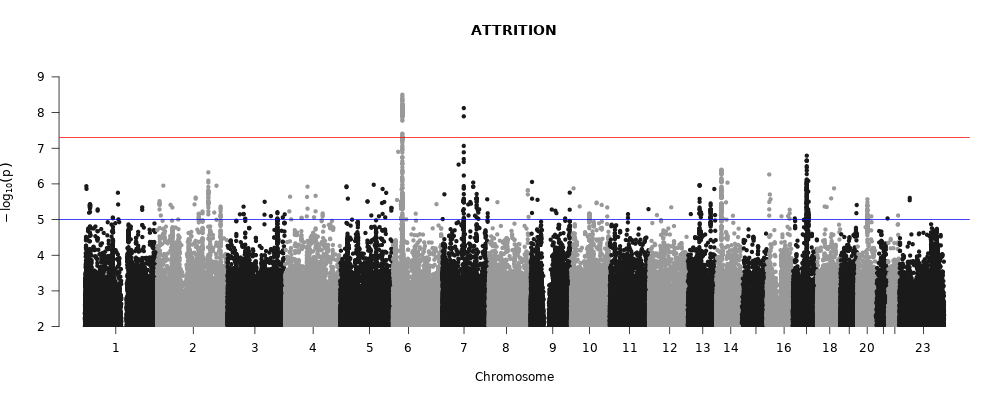

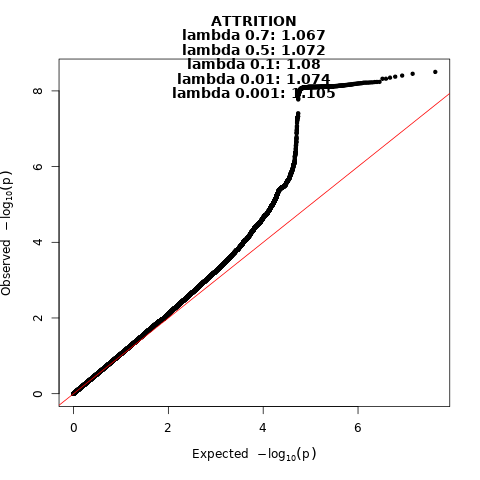

Figure 2. Manhattan and QQ-plot of phenotype Attrition.

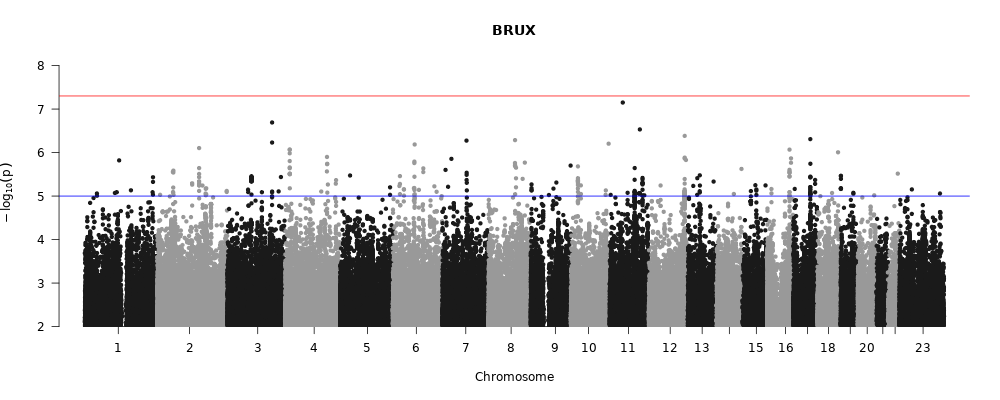

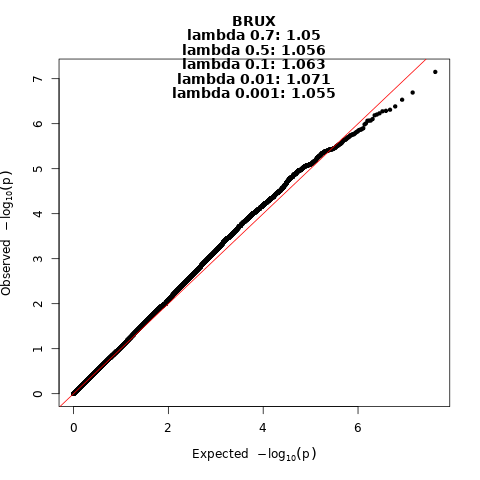

Figure 3. Manhattan and QQ-plot of phenotype Bruxism.

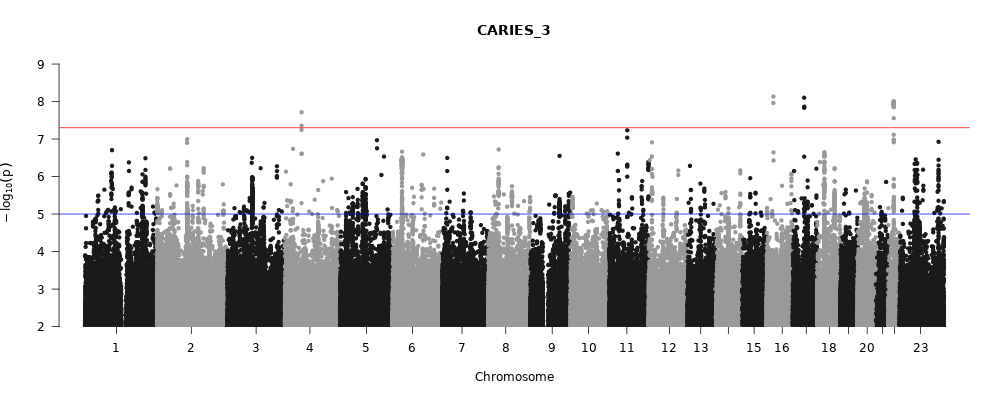

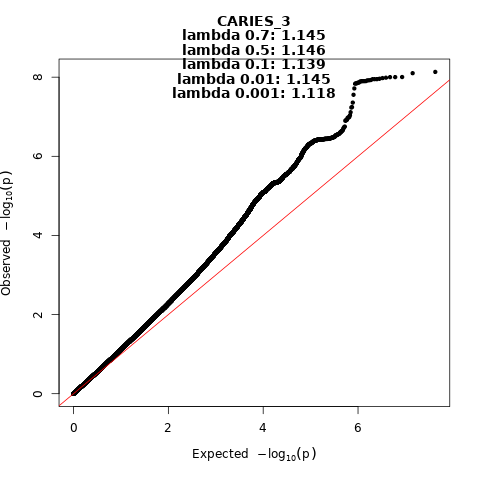

Figure 4. Manhattan and QQ-plot of phenotype Caries.

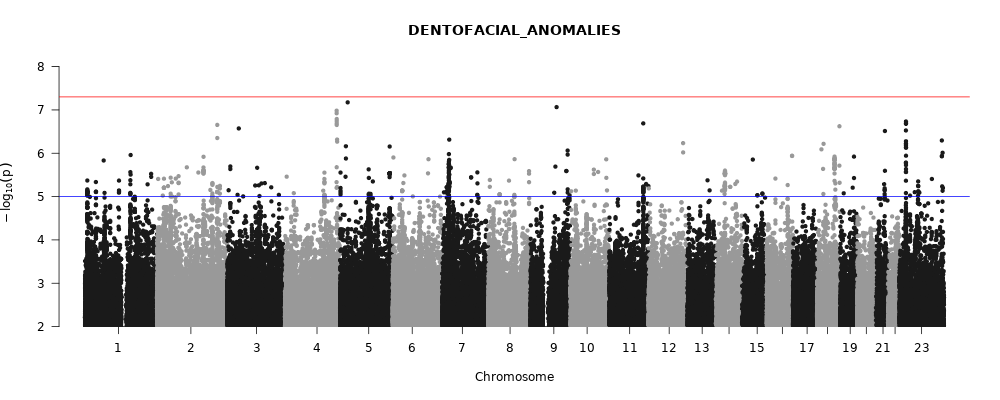

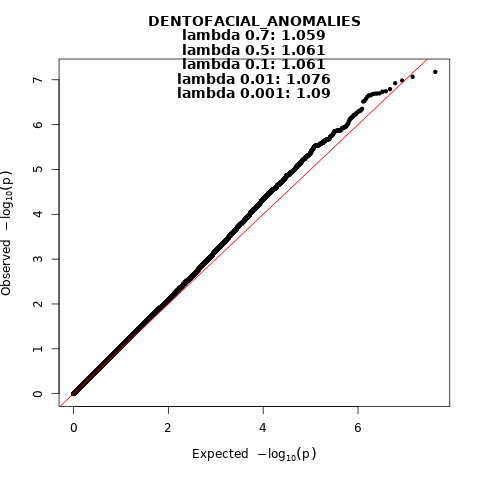
 Figure 5. Manhattan and QQ-plot of phenotype Dentofacial anomalies.

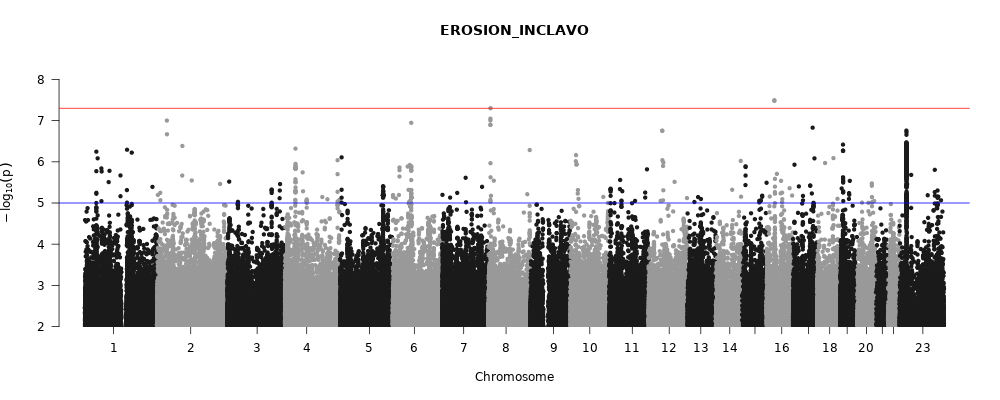

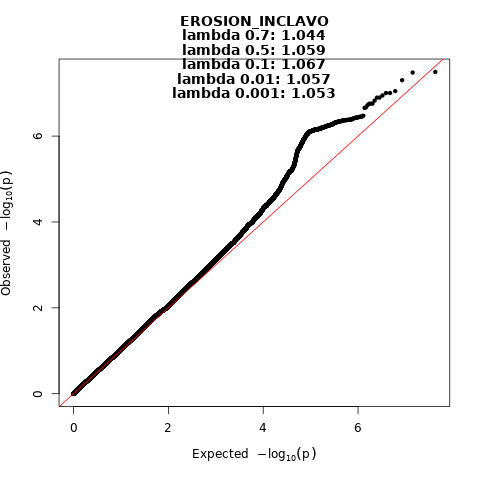
 Figure 6. Manhattan and QQ-plot of phenotype Erosion.

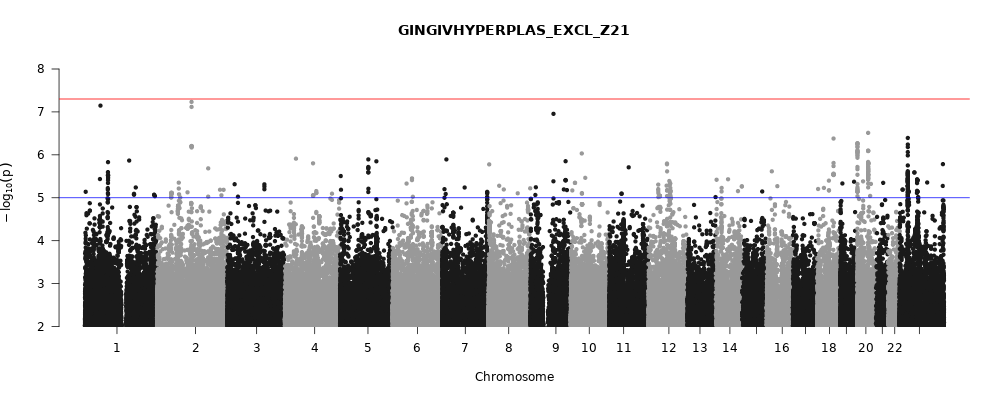

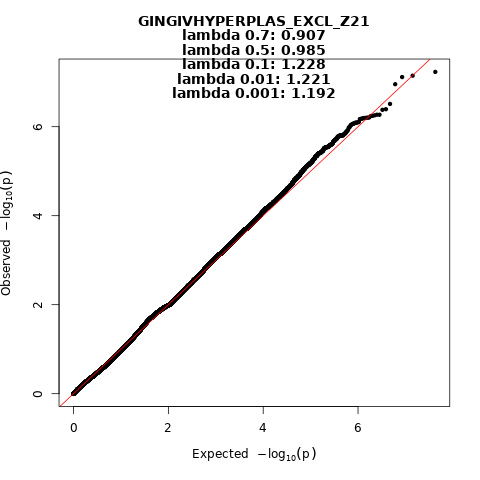

Figure 7. Manhattan and QQ-plot of phenotype Gingival hyperplasia.

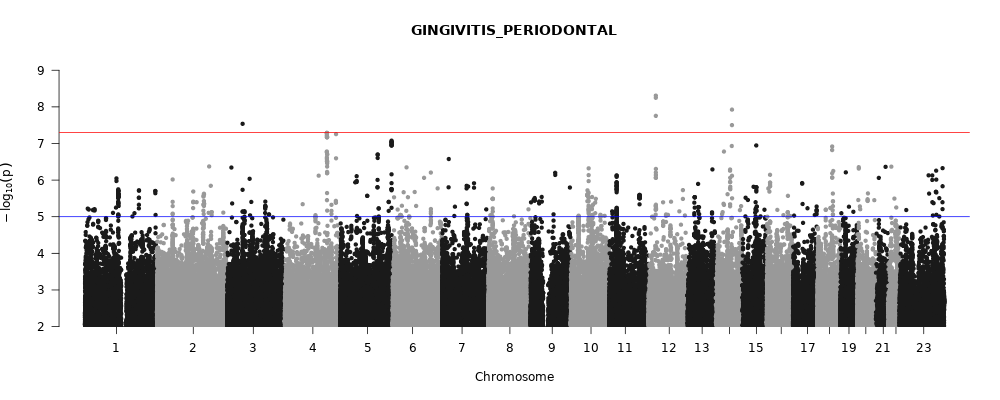

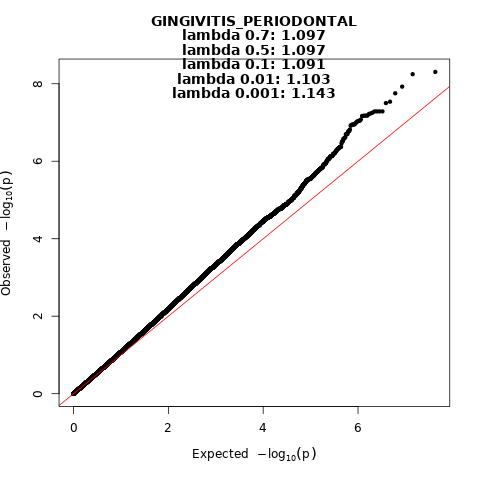
 Figure 8. Manhattan and QQ-plot of phenotype Gingivitis and periodontal diseases.

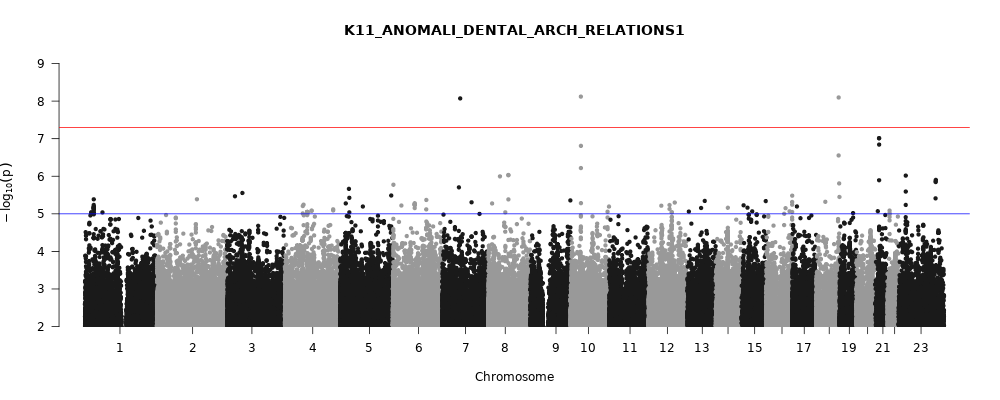

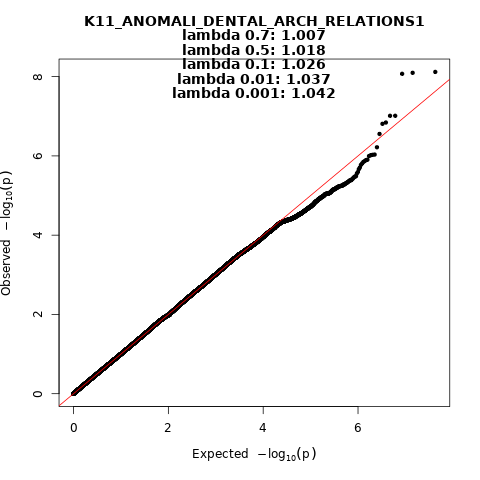
 Figure 9. Manhattan and QQ-plot of phenotype Anomalies of dental arch relationship.

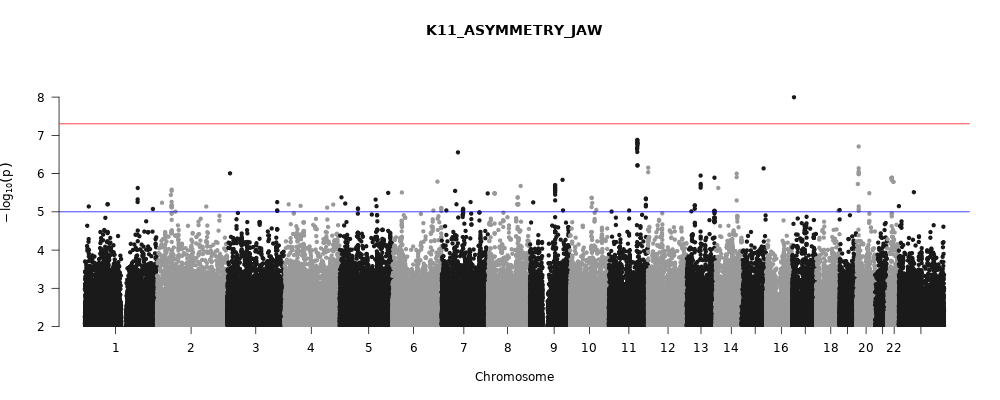

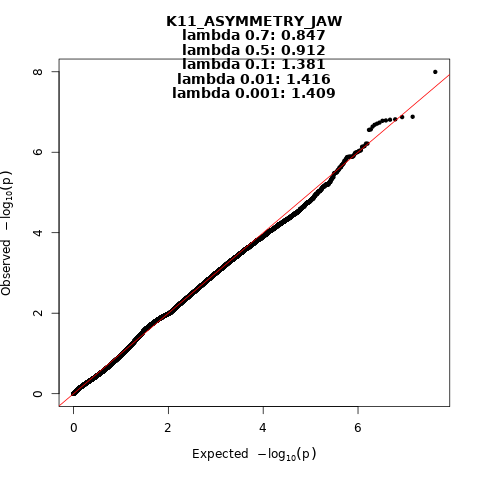
 Figure 10. Manhattan and QQ-plot of phenotype Asymmetry of jaw.

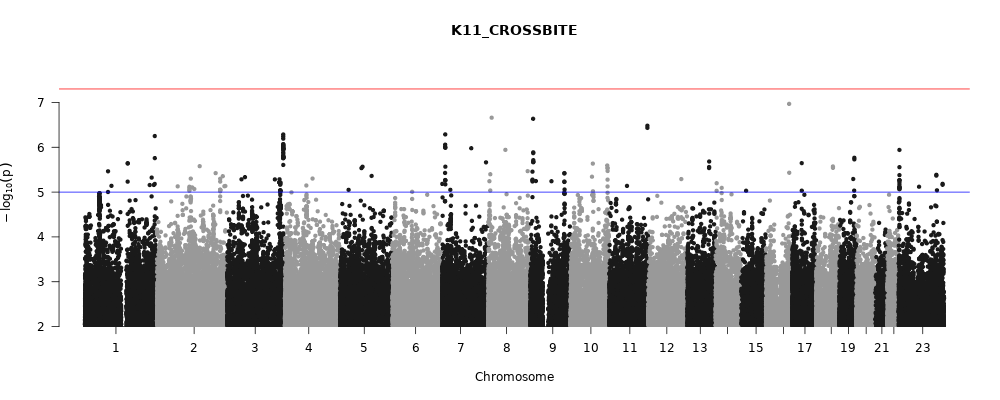

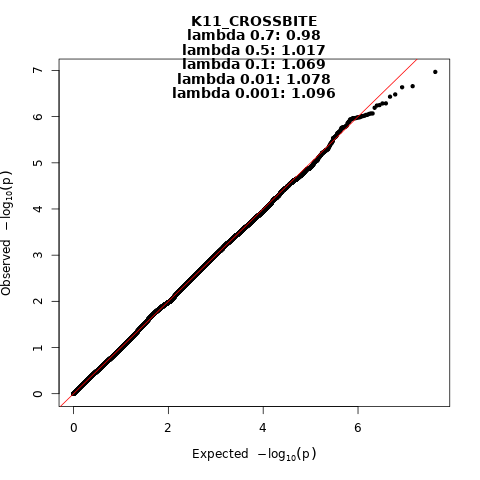
 Figure 11. Manhattan and QQ-plot of phenotype Crossbite.

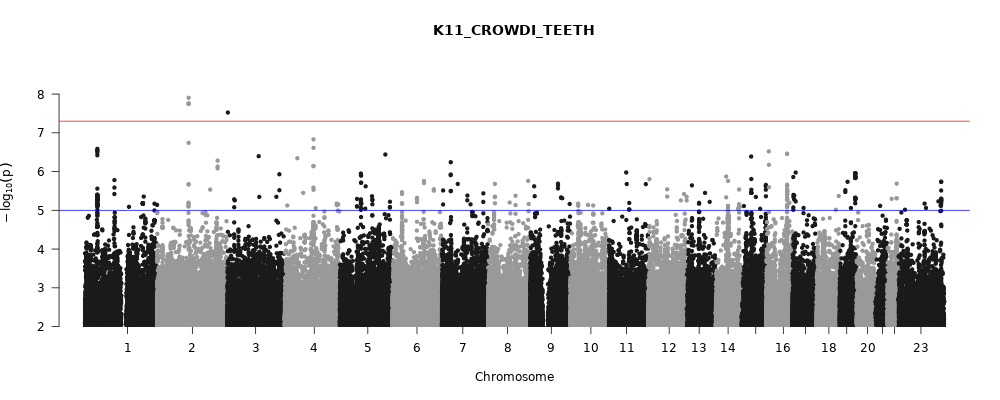

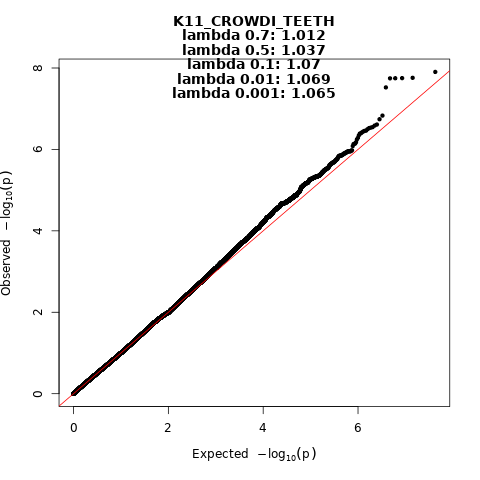
 Figure 12. Manhattan and QQ-plot of phenotype Crowding of teeth.

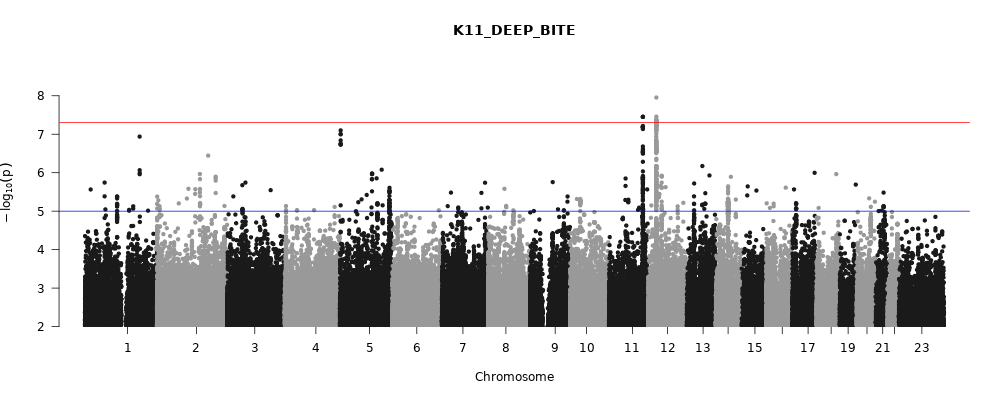

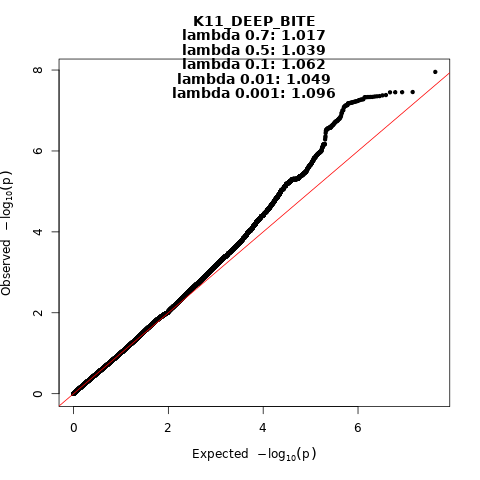
 Figure 13. Manhattan and QQ-plot of phenotype Deep bite.

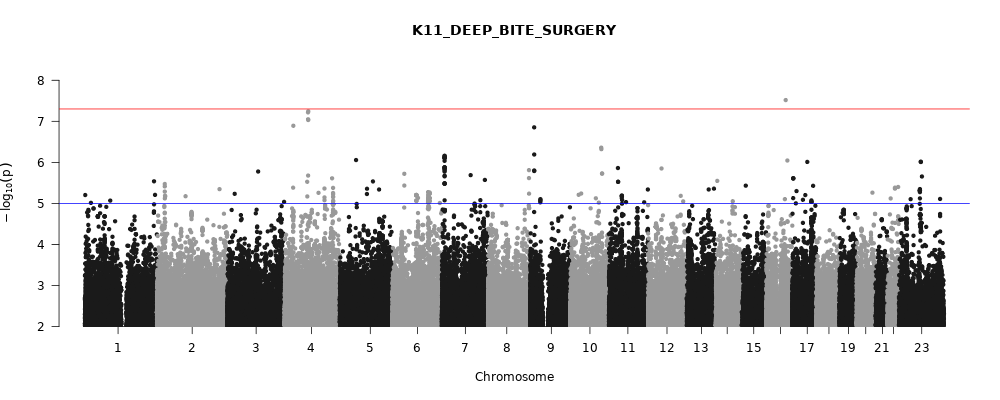

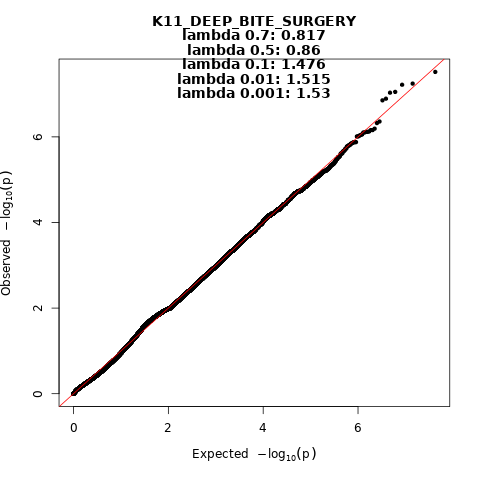
 Figure 14. Manhattan and QQ-plot of phenotype Deep bite that required surgery.

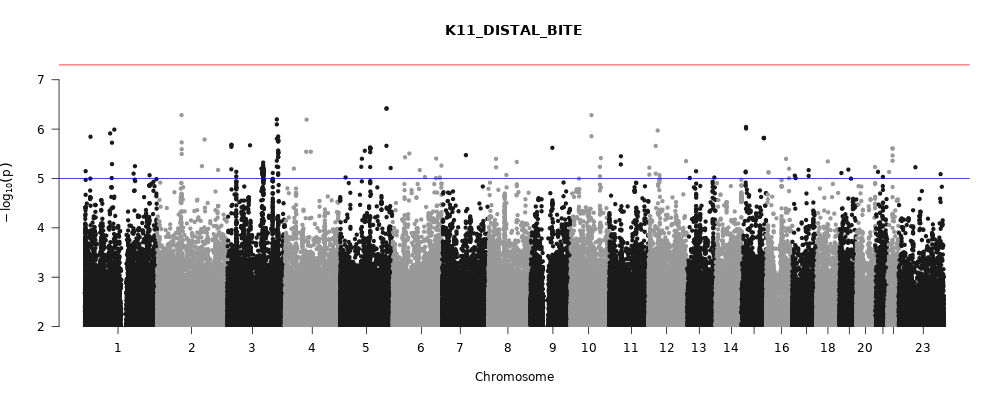

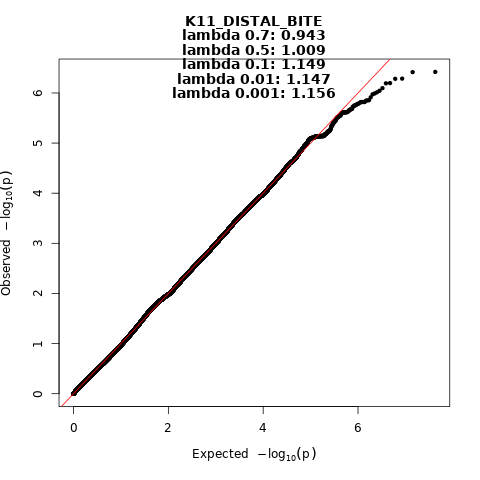
 Figure 15. Manhattan and QQ-plot of phenotype Disto-occlusion.

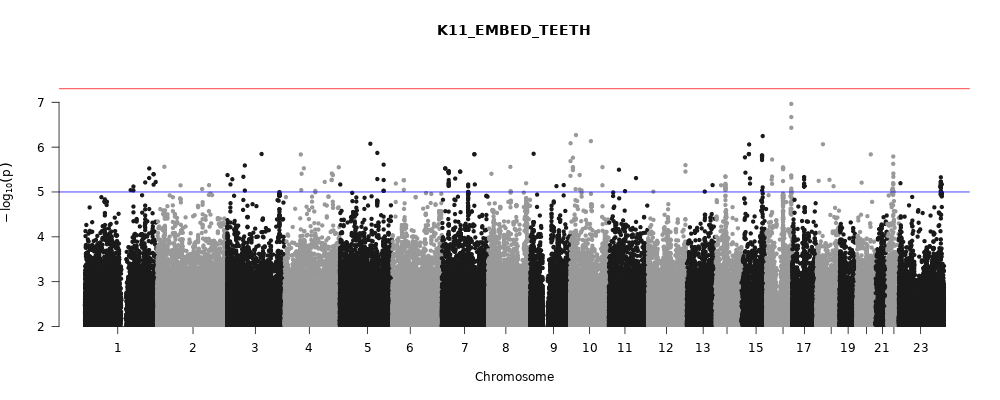

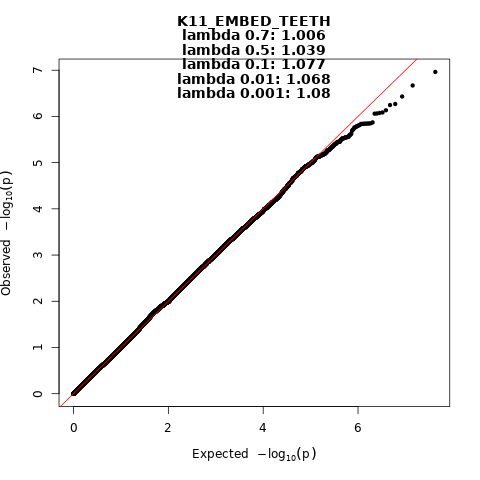
 Figure 16. Manhattan and QQ-plot of phenotype Embedded teeth.

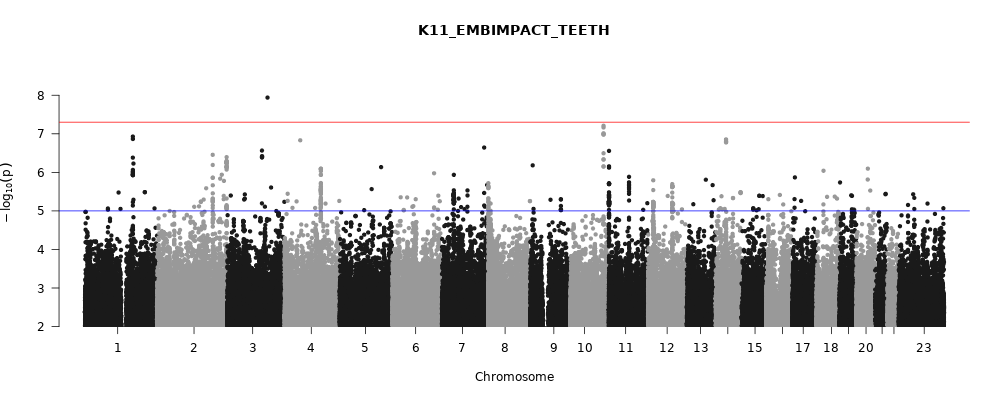

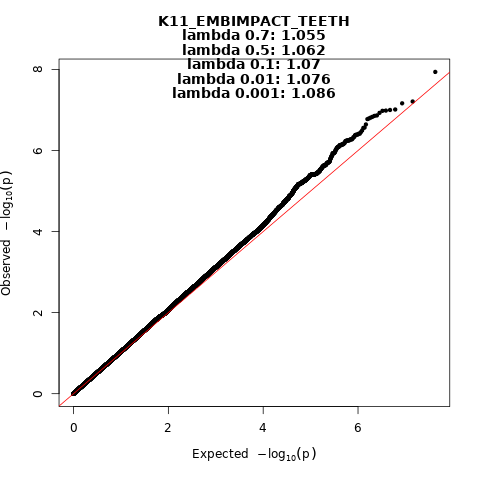
 Figure 17. Manhattan and QQ-plot of phenotype Embedded and impacted teeth.

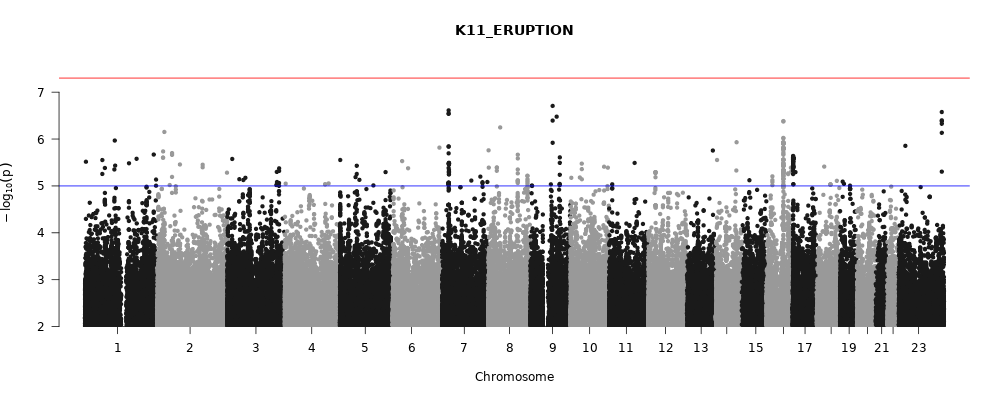

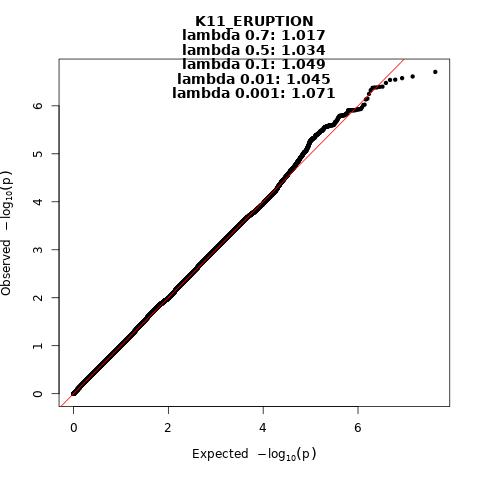
 Figure 18. Manhattan and QQ-plot of phenotype Tooth eruption problems.

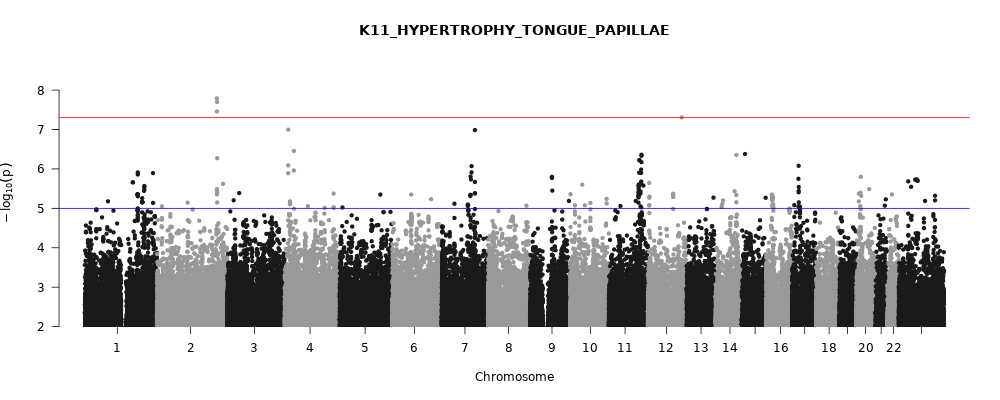

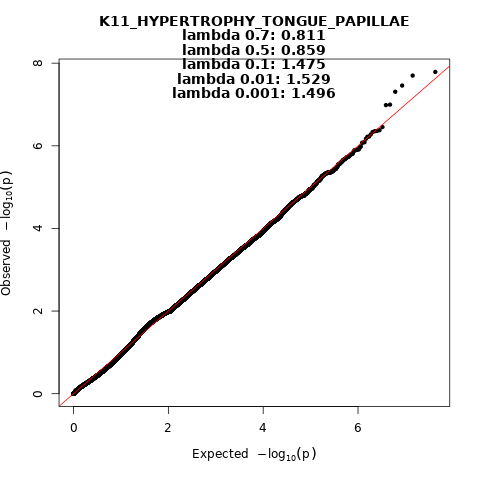
 Figure 19. Manhattan and QQ-plot of phenotype Hypertrophy of tongue papillae.

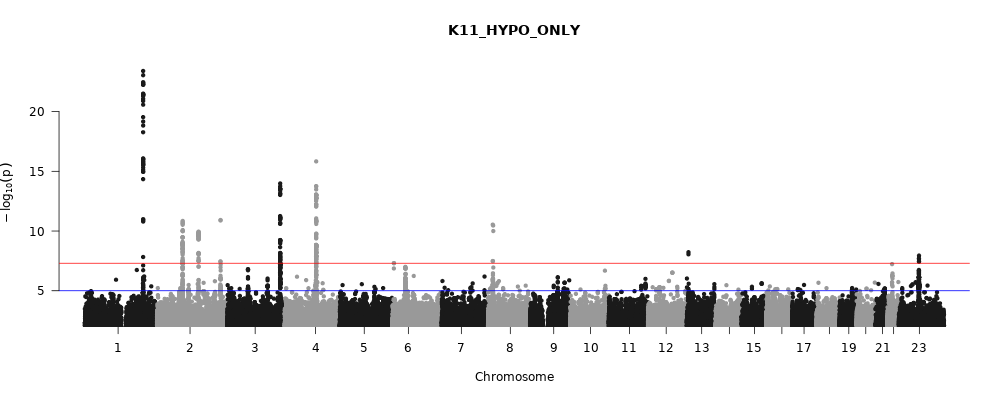

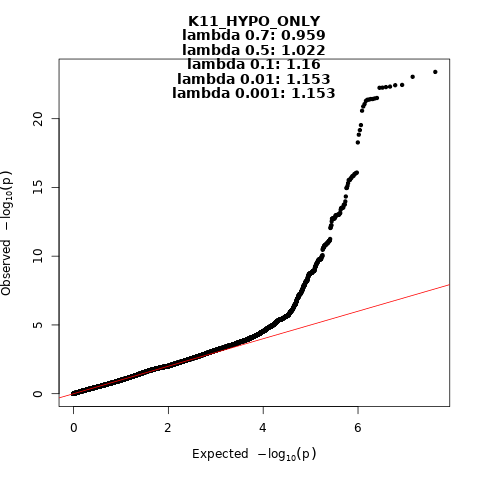
 Figure 20. Manhattan and QQ-plot of phenotype Hypodontia or oligodontia.

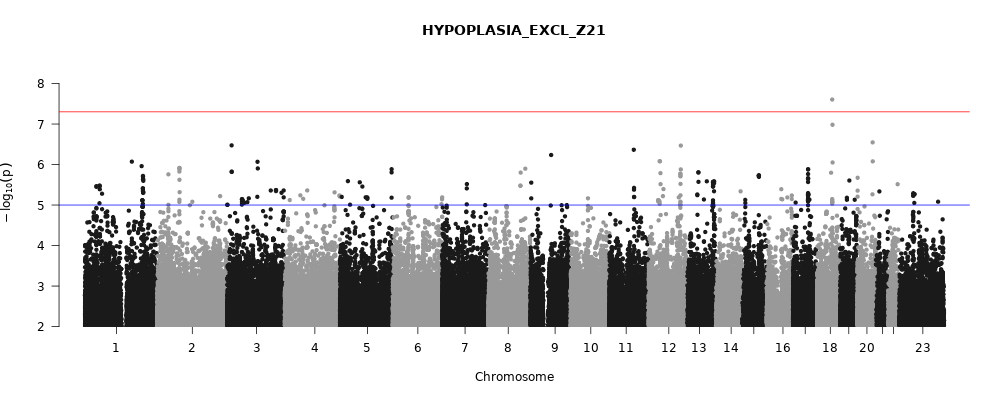

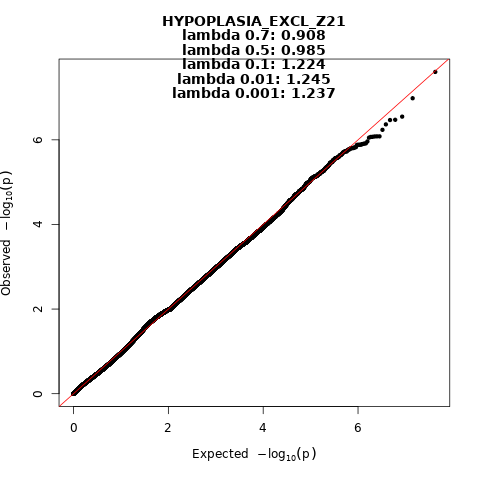

Figure 21. Manhattan and QQ-plot of phenotype Hypoplasia of dental enamel.
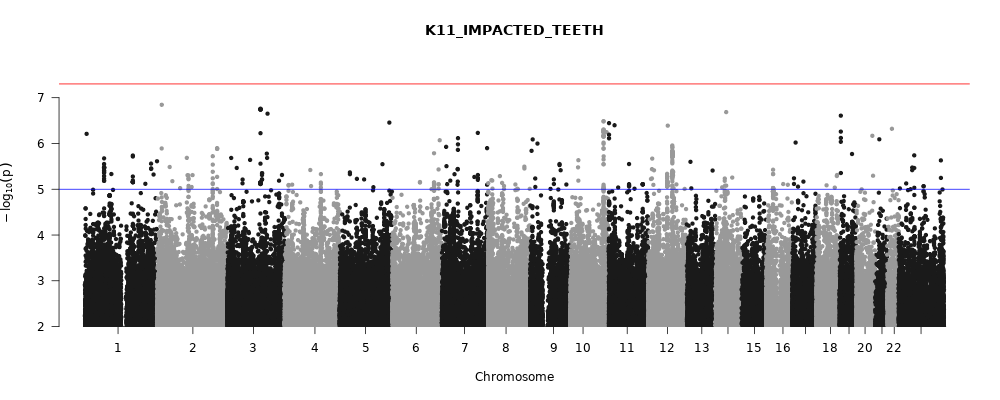

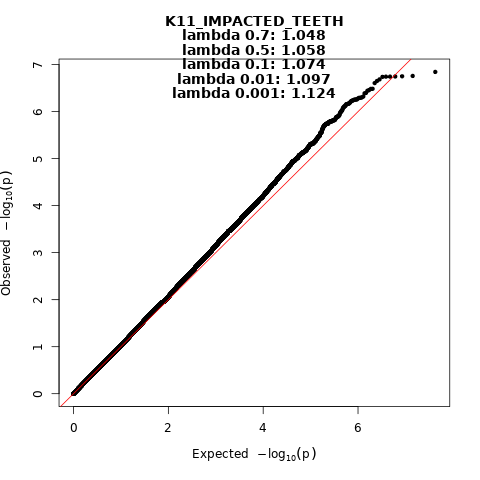
 Figure 22. Manhattan and QQ-plot of phenotype Impacted teeth.
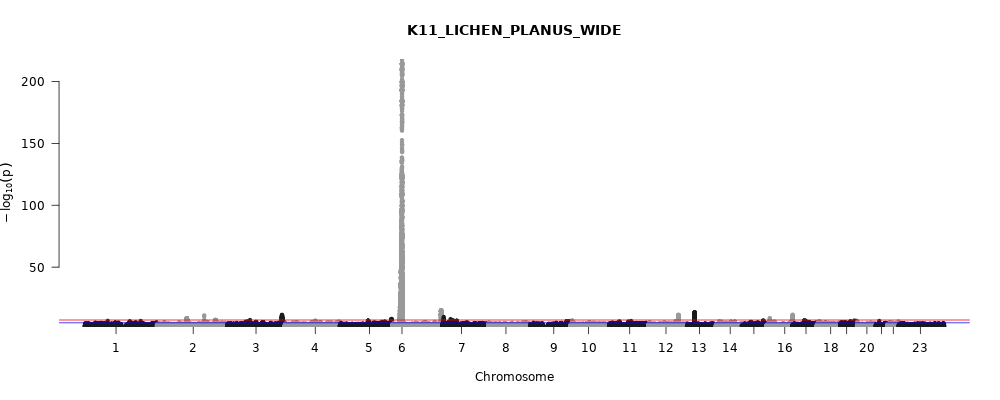

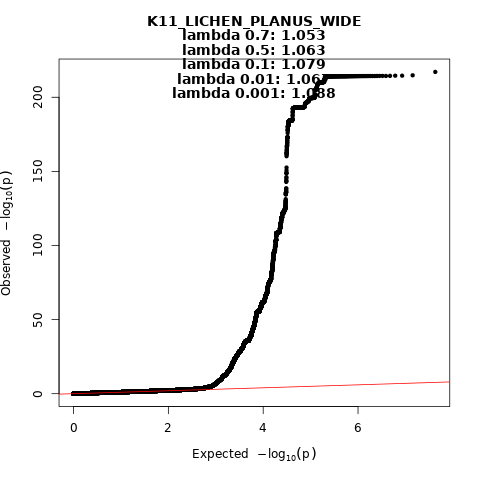
 Figure 23. Manhattan and QQ-plot of phenotype Peripheral and oral lichen planus.
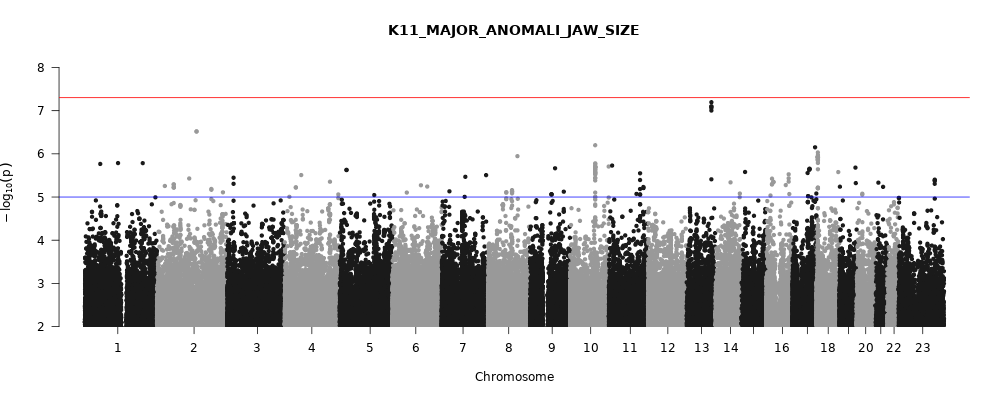

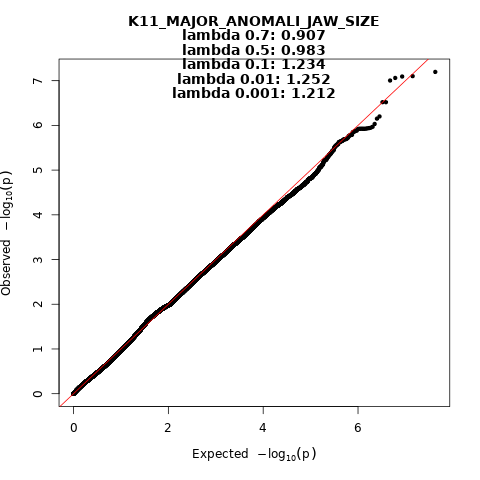
 Figure 24. Manhattan and QQ-plot of phenotype Major anomalies of jaw size.
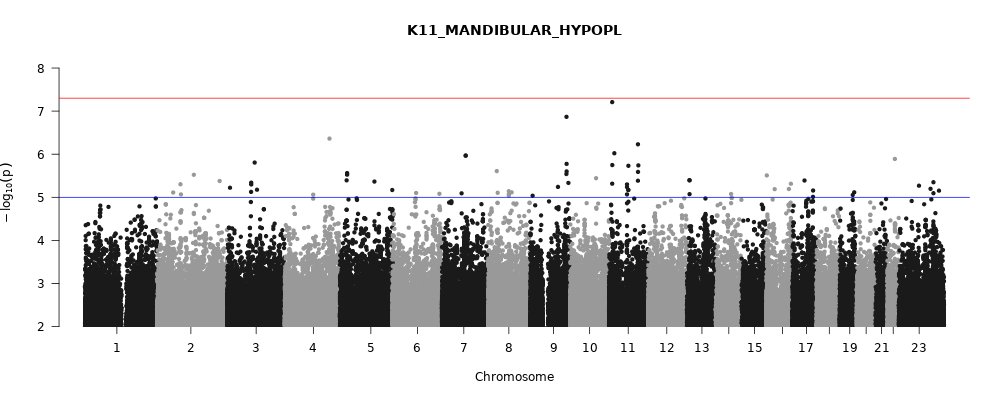

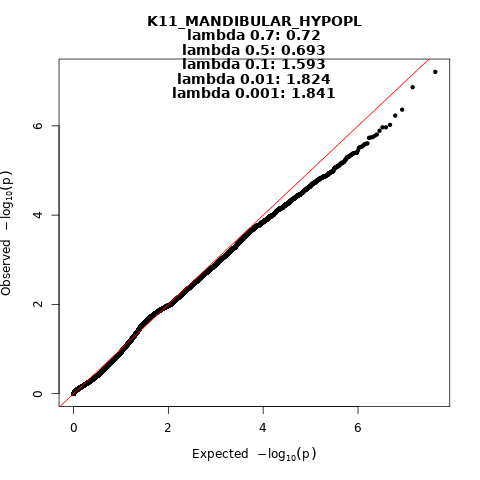
 Figure 25. Manhattan and QQ-plot of phenotype Mandibular hypoplasia.
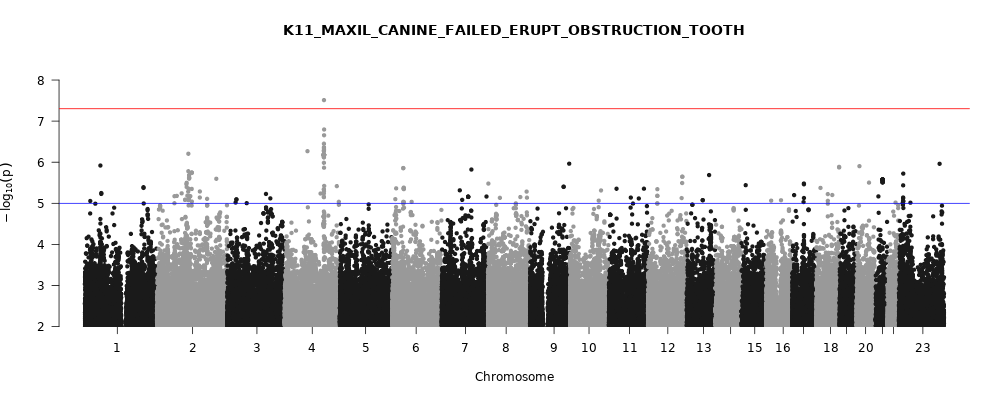

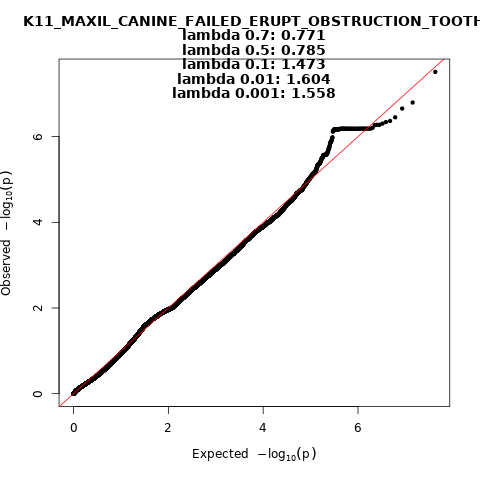
 Figure 26. Manhattan and QQ-plot of phenotype Impacted maxillary canine.
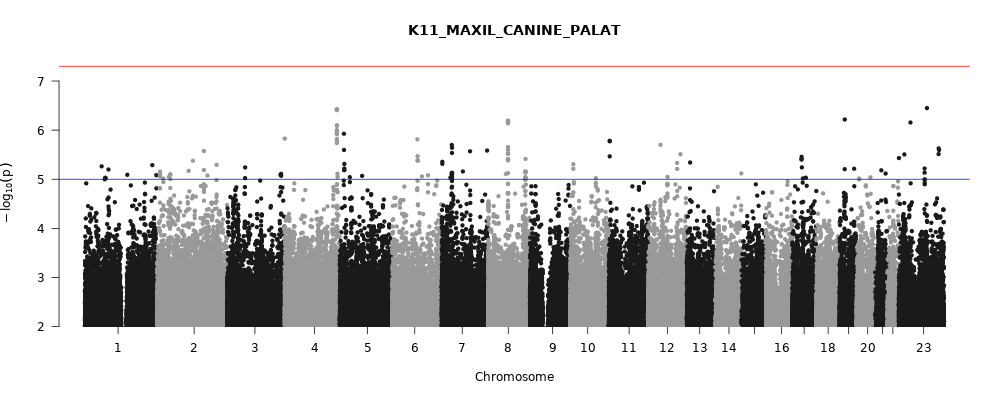

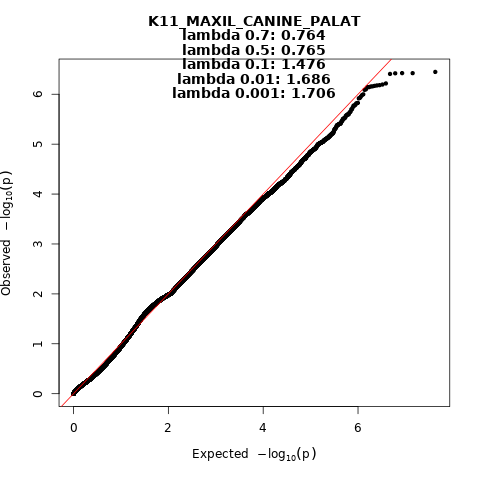
 Figure 27. Manhattan and QQ-plot of phenotype Palatinal maxillary canine.
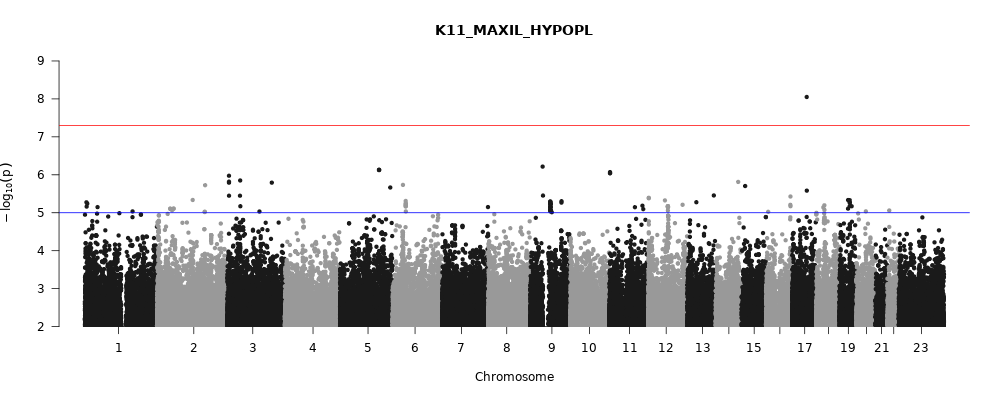

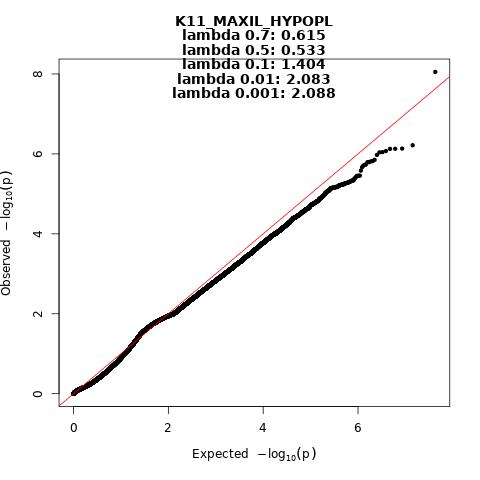
 Figure 28. Manhattan and QQ-plot of phenotype Maxillary hypoplasia.
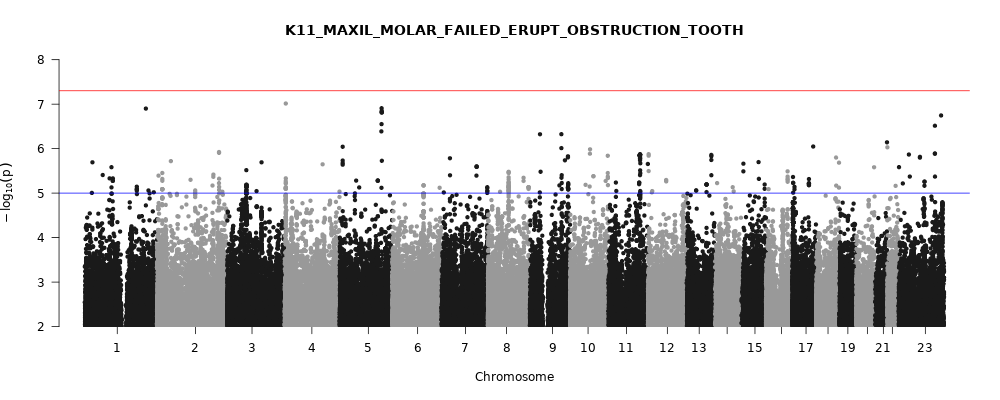

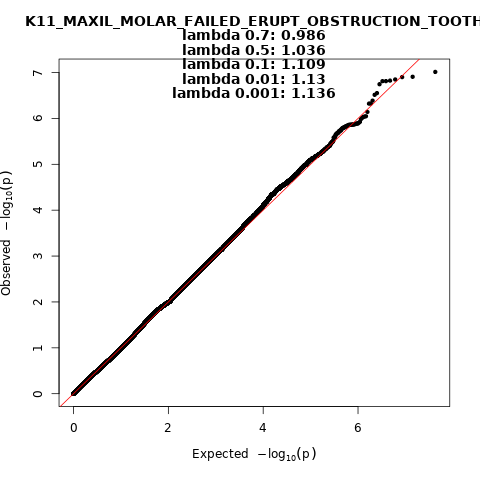
 Figure 29. Manhattan and QQ-plot of phenotype Impacted maxillary molar.
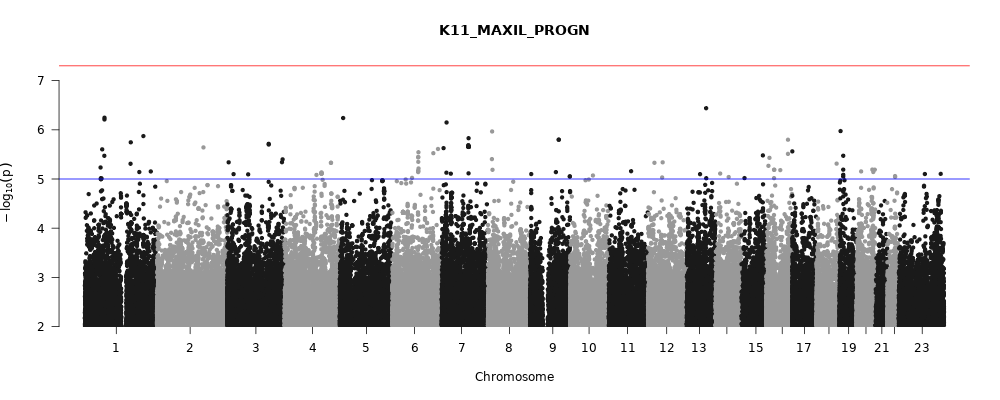

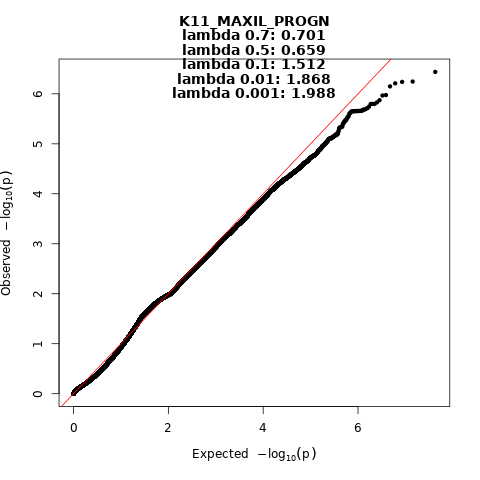
 Figure 30. Manhattan and QQ-plot of phenotype Maxillary prognathia.
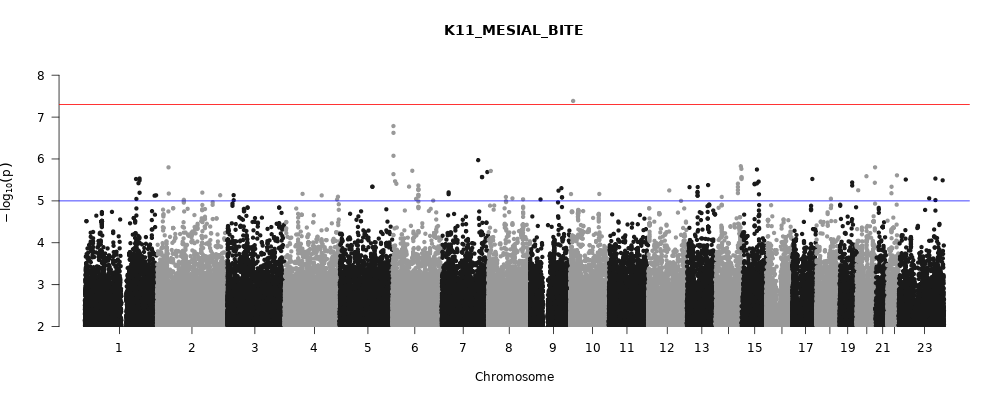

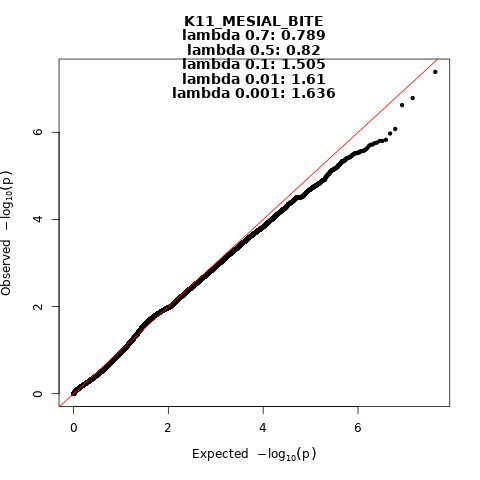
 Figure 31. Manhattan and QQ-plot of phenotype Mesio-occlusion.
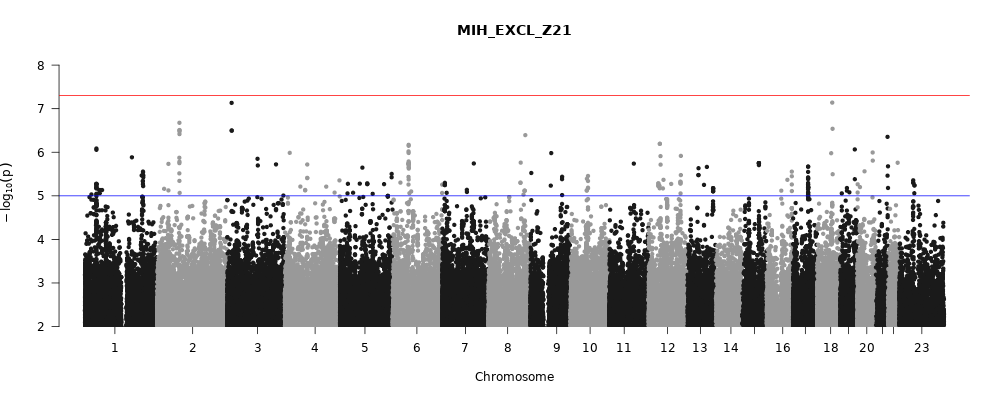

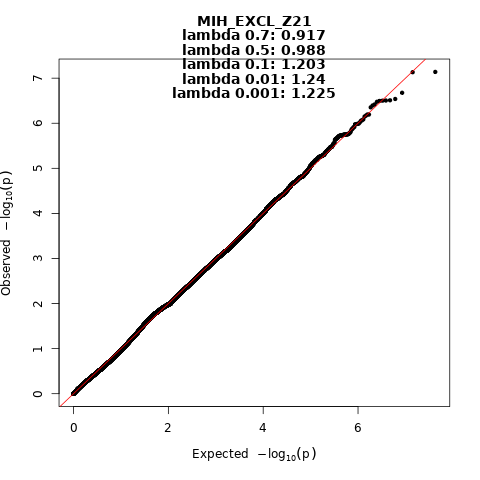
 Figure 32. Manhattan and QQ-plot of phenotype Molar incisor hypomineralisation (MIH).
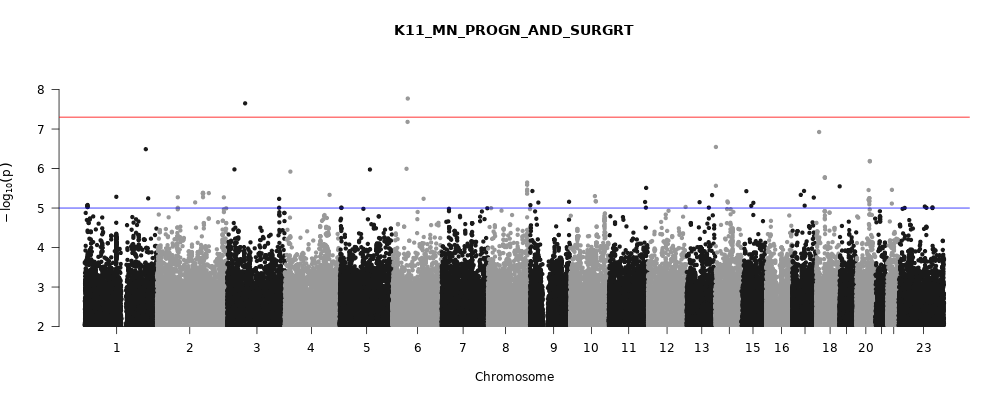

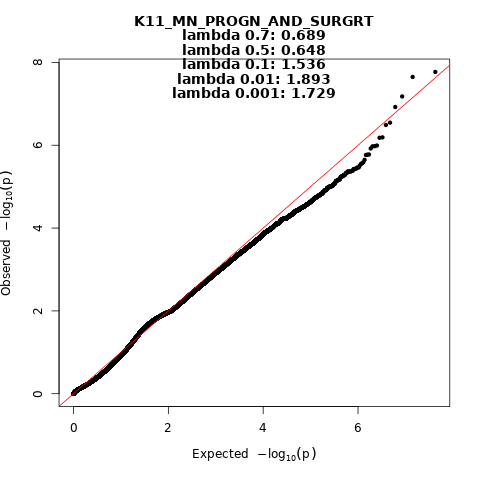
 Figure 33. Manhattan and QQ-plot of phenotype Mandibular prognathia and surgery (LeFortI or BSSRO).
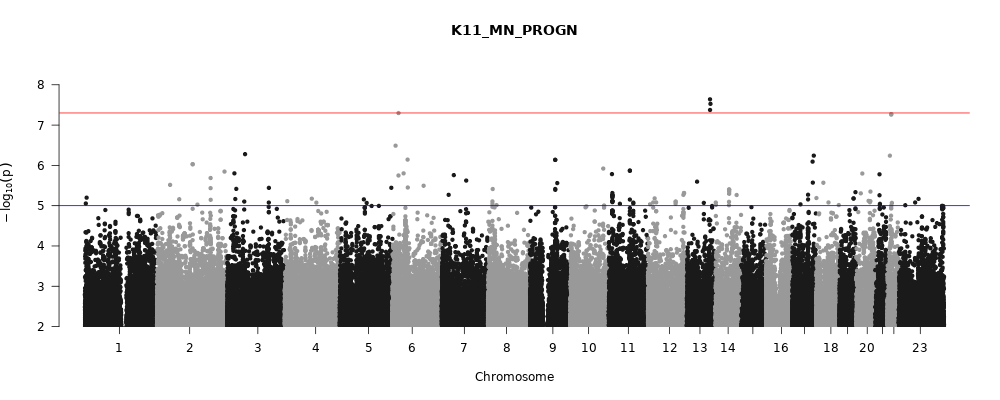

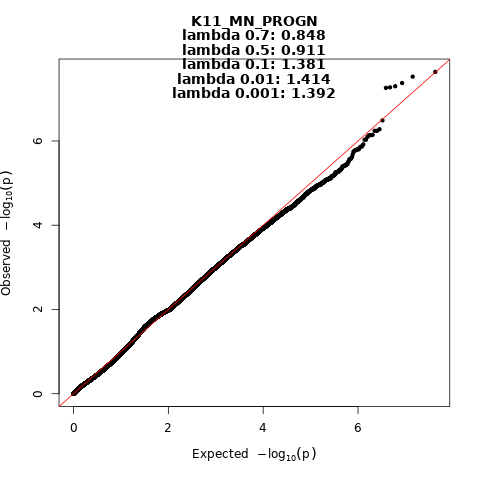
 Figure 34. Manhattan and QQ-plot of phenotype Mandibular prognathia.
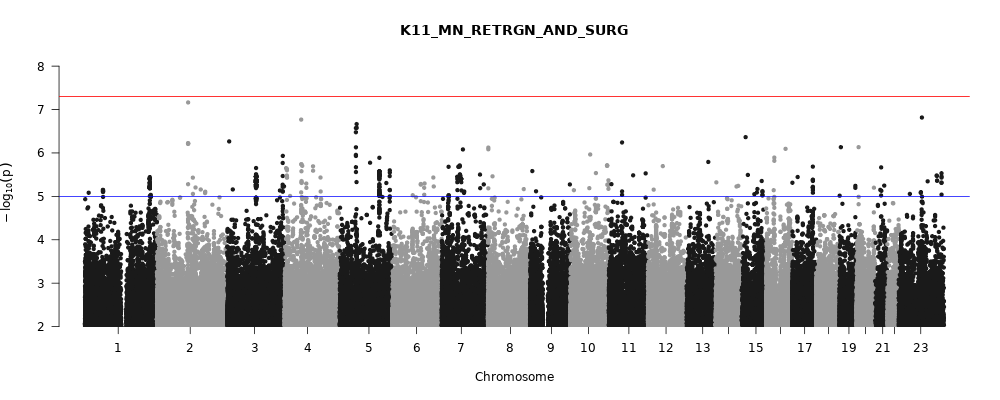

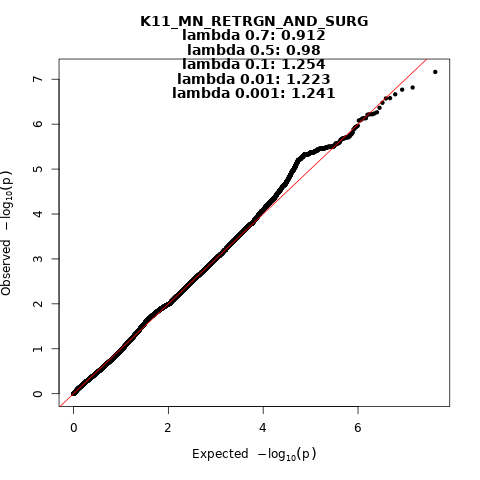
 Figure 35. Manhattan and QQ-plot of phenotype Mandibular retrognathia and surgery (BSSRO).
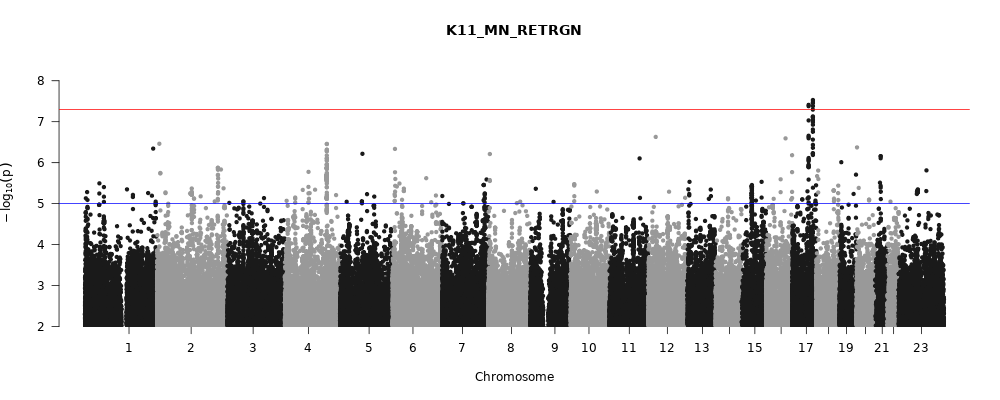

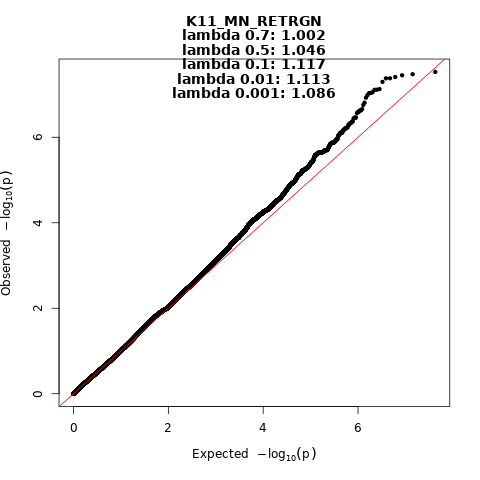
 Figure 36. Manhattan and QQ-plot of phenotype Mandibular retrognathia.
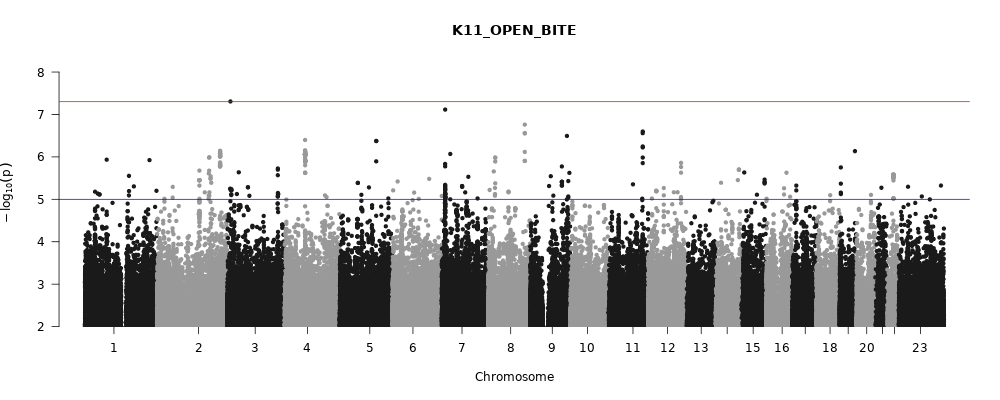

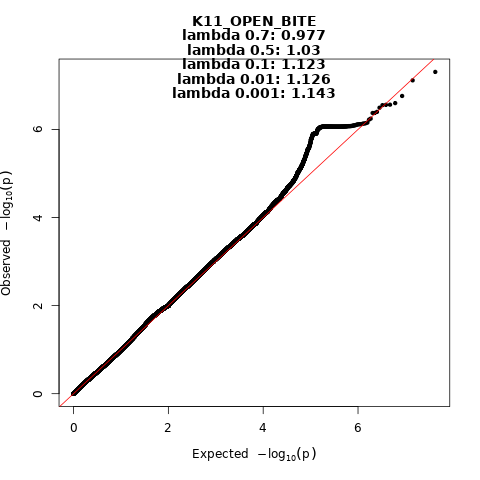
 Figure 37. Manhattan and QQ-plot of phenotype Open bite.
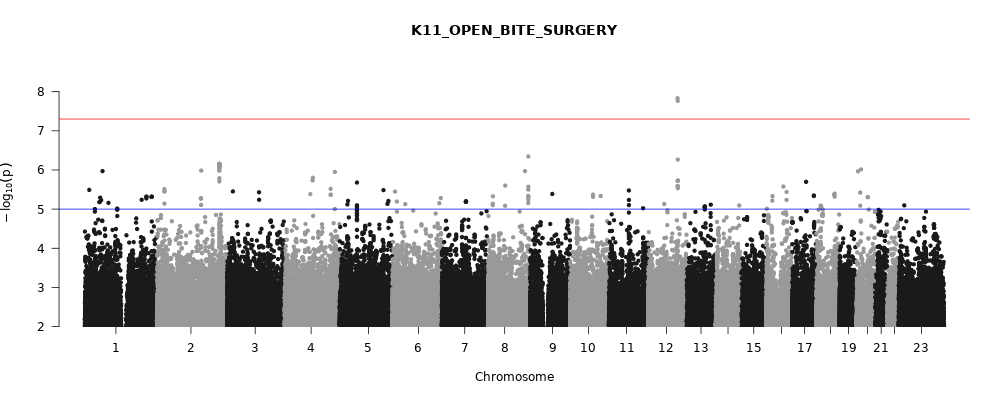

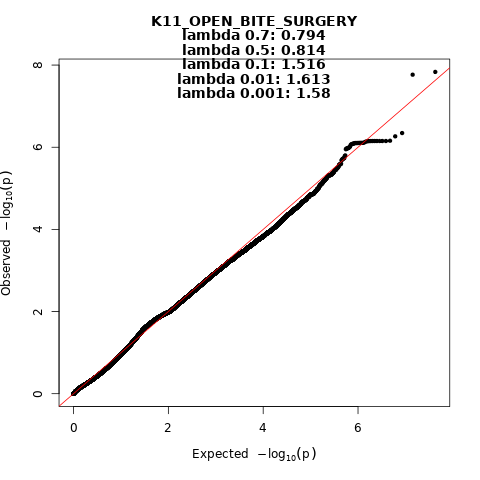
 Figure 38. Manhattan and QQ-plot of phenotype Open bite that required surgery (LeFort1 or BSSRO).

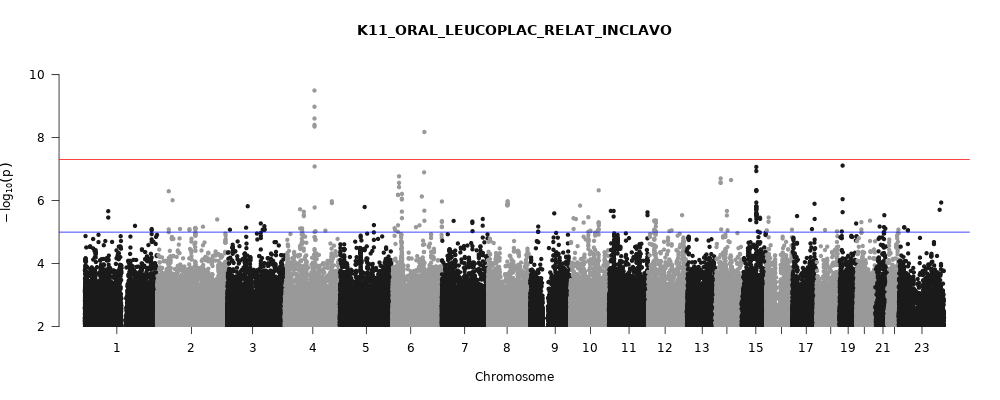

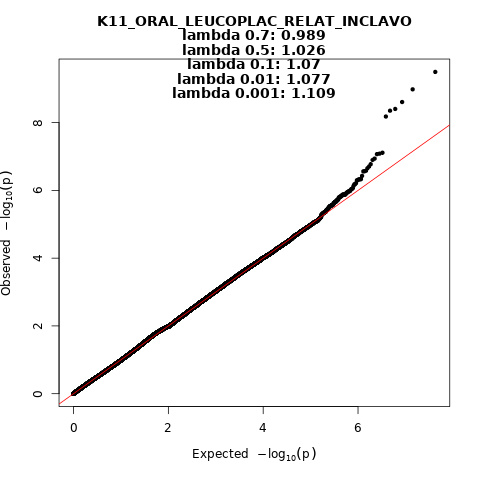
 Figure 39. Manhattan and QQ-plot of phenotype Oral leukoplakia and related diseases.
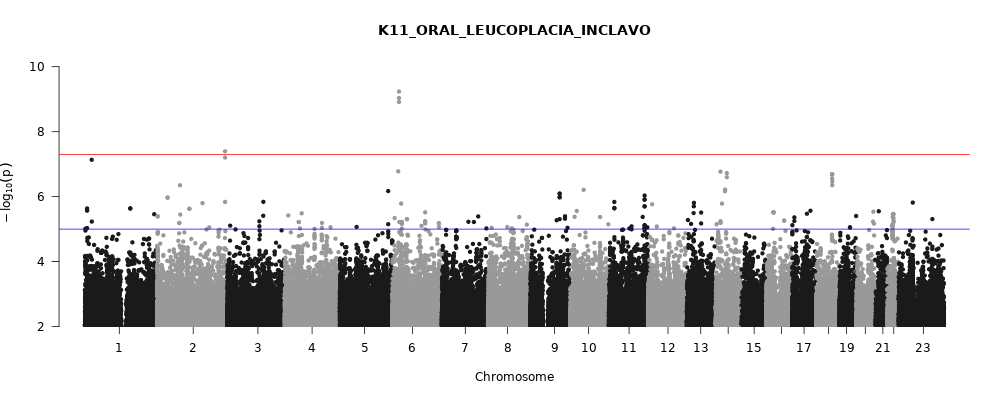

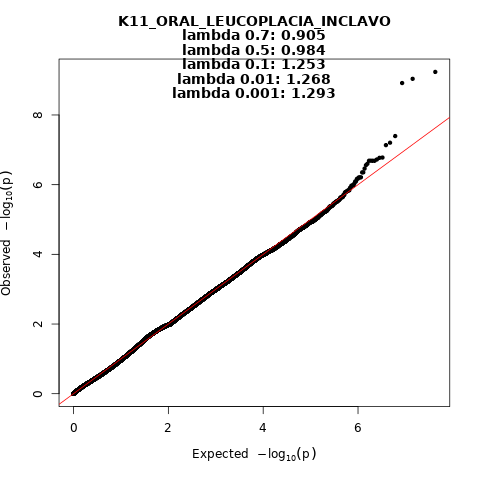
 Figure 40. Manhattan and QQ-plot of phenotype Oral leukoplakia.
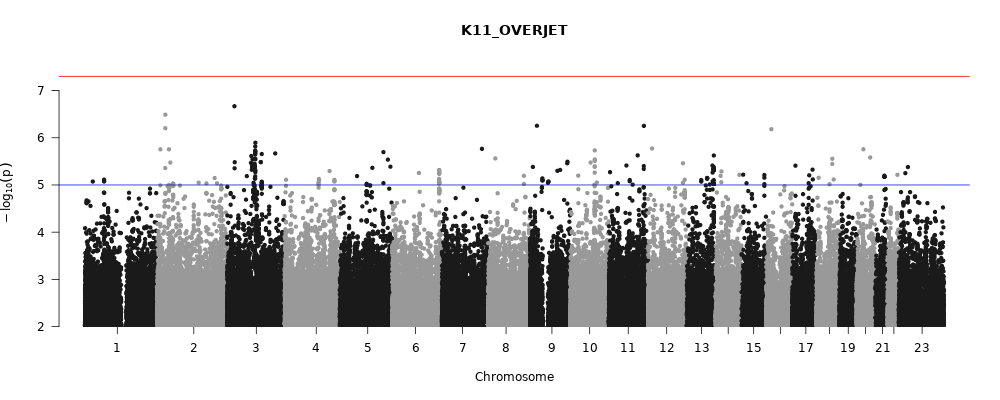

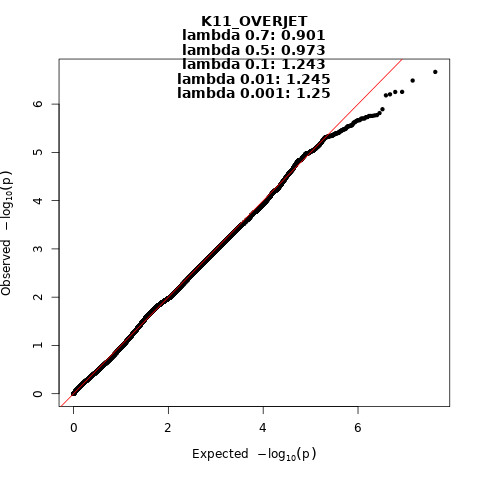
 Figure 41. Manhattan and QQ-plot of phenotype Overjet.
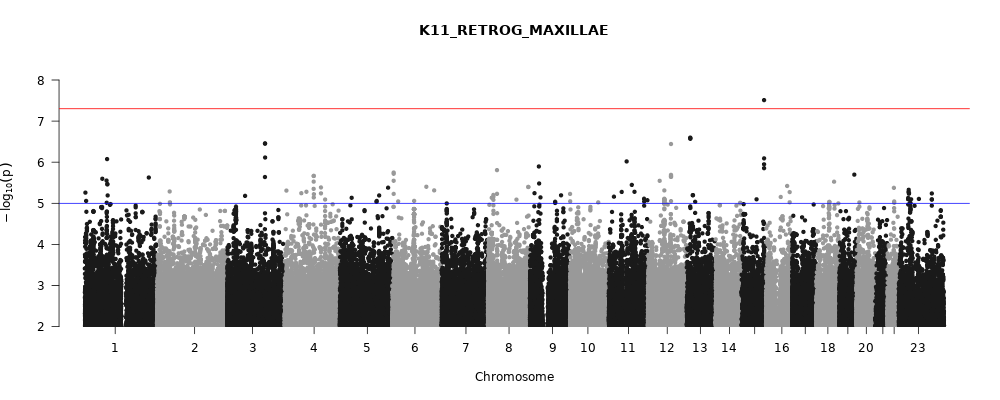

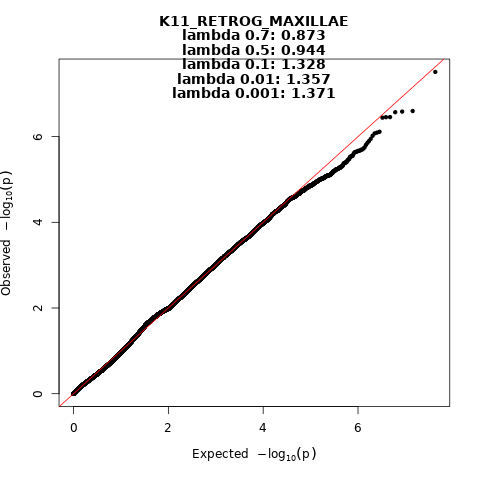
 Figure 42. Manhattan and QQ-plot of phenotype Maxillary retrognathism.
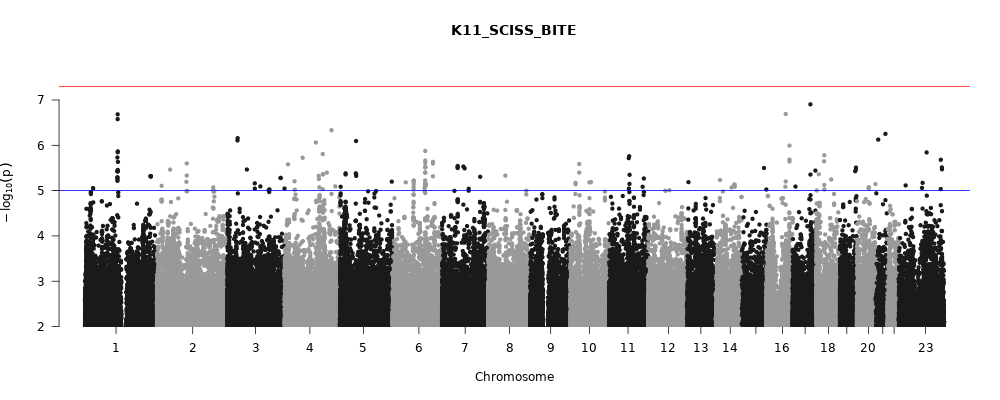

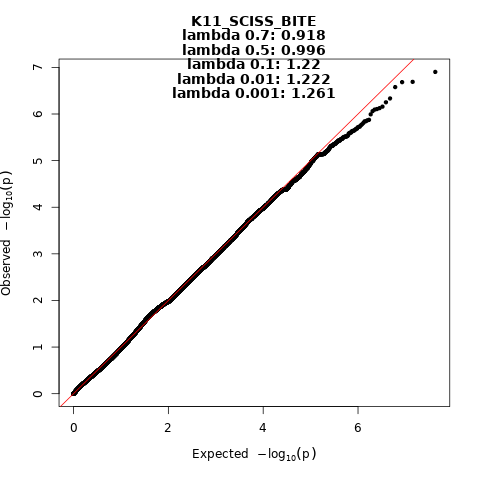
 Figure 43. Manhattan and QQ-plot of phenotype Scissor bite.
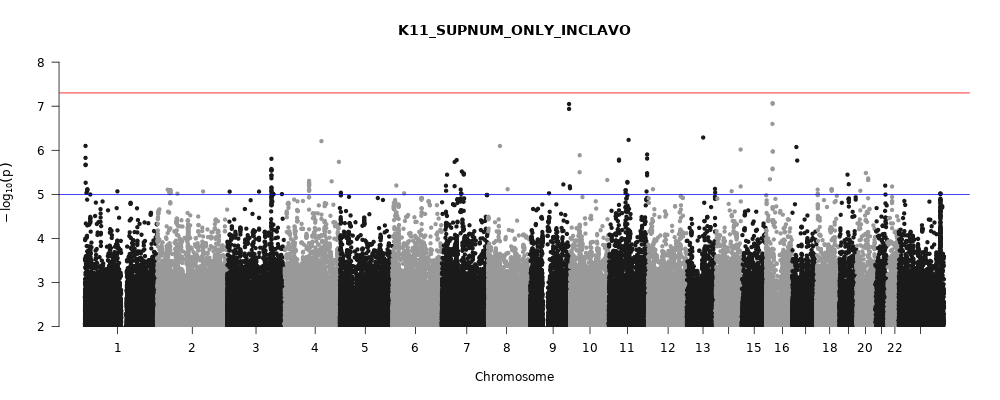

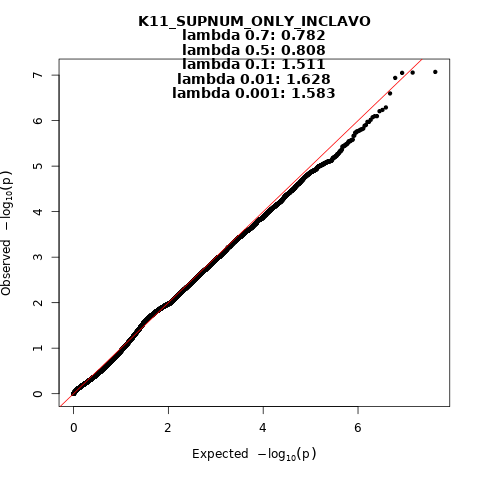
 Figure 44. Manhattan and QQ-plot of phenotype Supernumerary teeth (excluding clefts and syndromes).
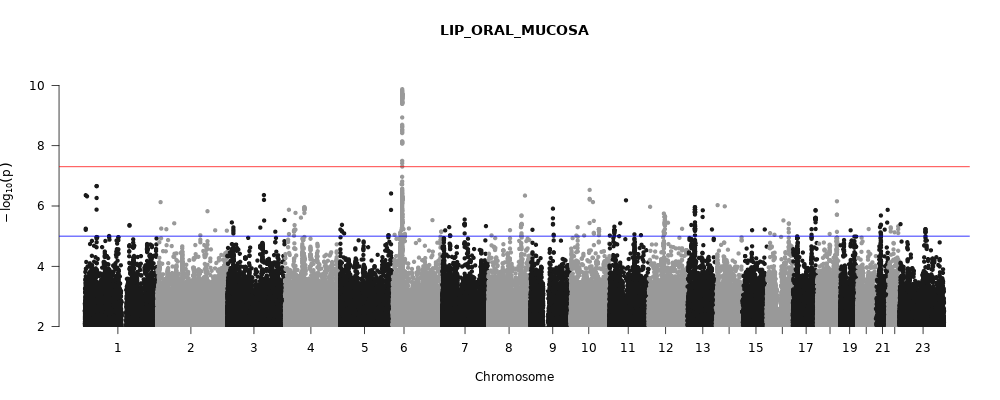

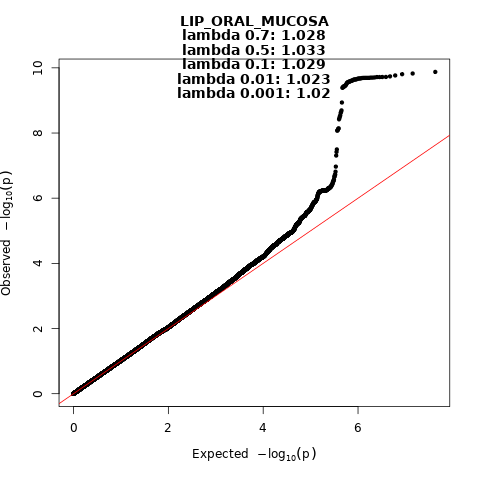
 Figure 45. Manhattan and QQ-plot of phenotype Diseases of lip and oral mucosa.
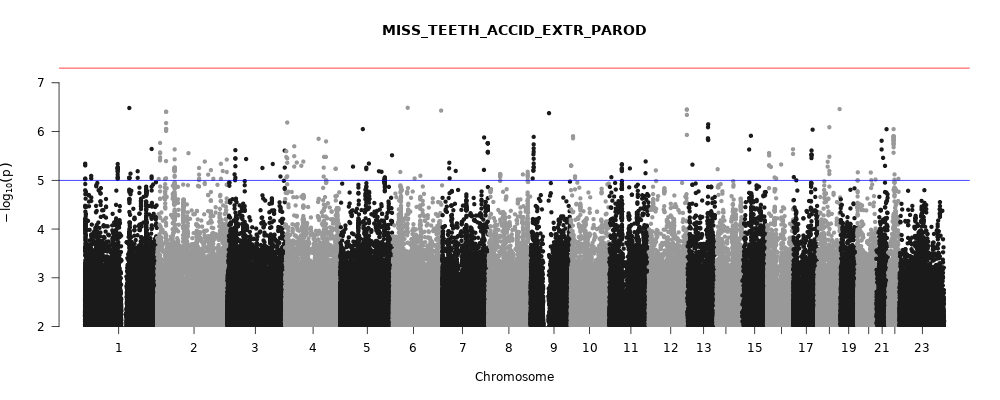

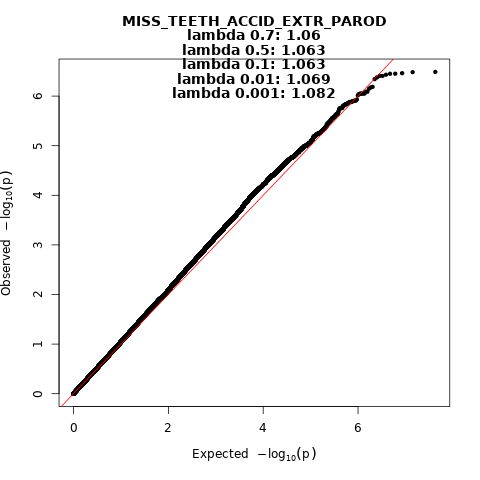
 Figure 46. Manhattan and QQ-plot of phenotype Tooth loss.
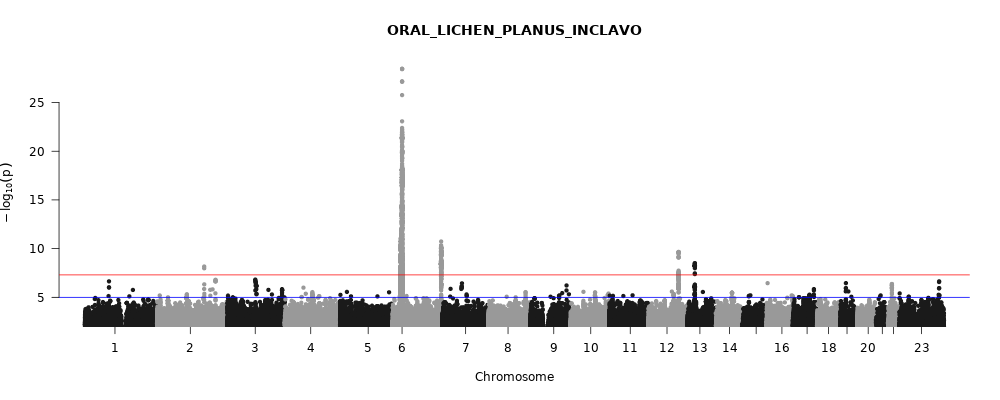

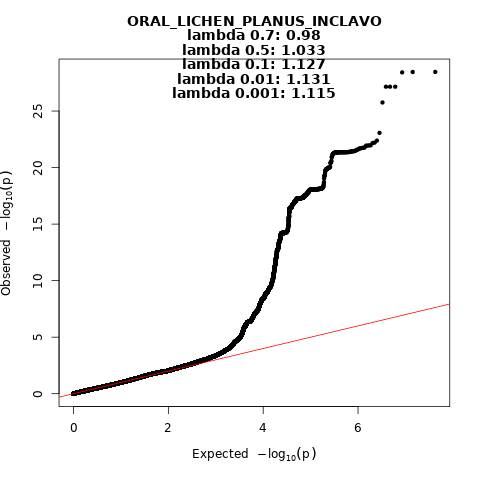
 Figure 47. Manhattan and QQ-plot of phenotype Oral lichen planus.
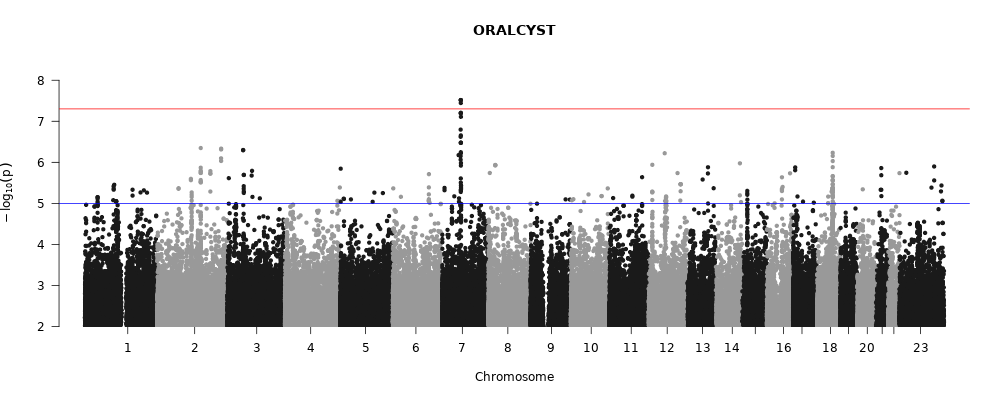

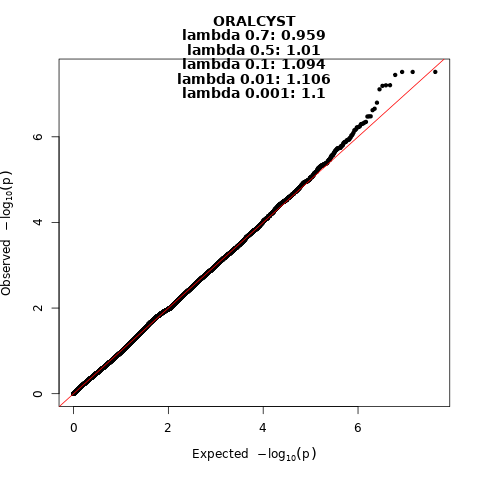
 Figure 48. Manhattan and QQ-plot of phenotype Oral cysts.
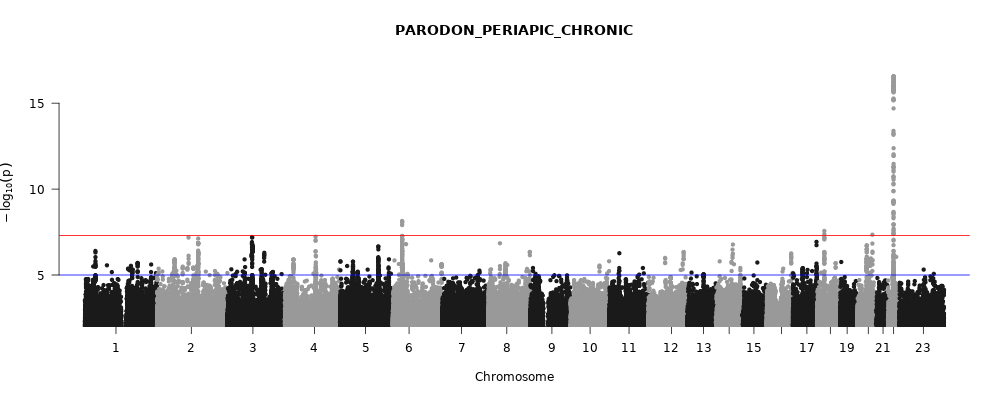

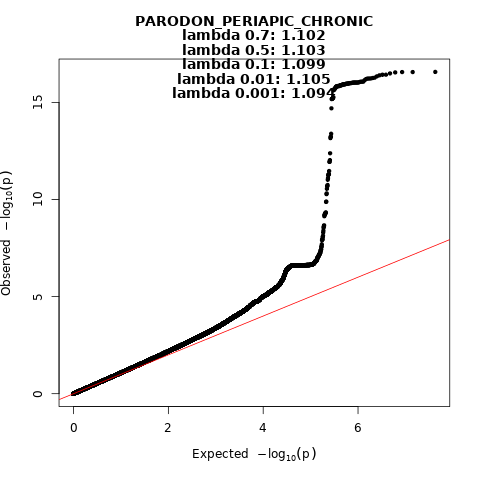
 Figure 49. Manhattan and QQ-plot of phenotype Chronic apical periodontitis.
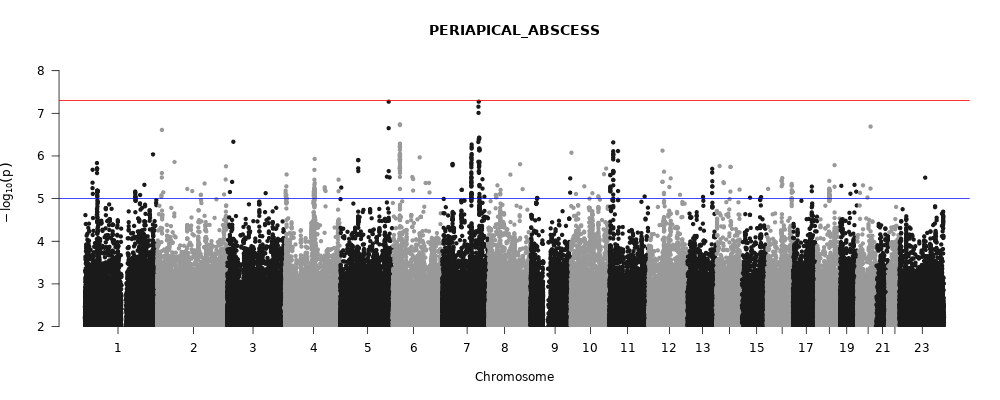

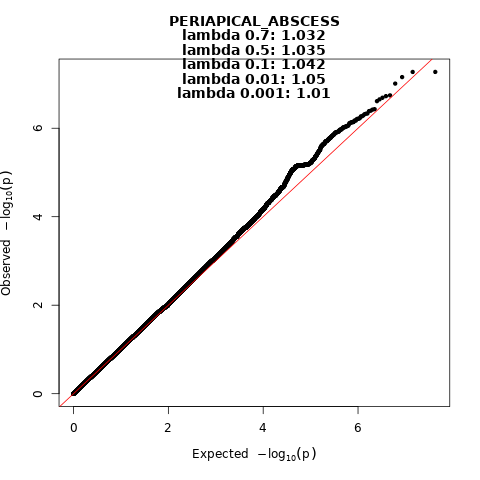
 Figure 50. Manhattan and QQ-plot of phenotype Periapical abscess. Figure 51. Manhattan and QQ-plot of phenotype Chronic complicated periodontitis. Figure 52. Manhattan and QQ-plot of phenotype Chronic periodontitis. Figure 53. Manhattan and QQ-plot of phenotype Aggressive periodontitis. Figure 54. Manhattan and QQ-plot of phenotype Necrosis of pulp. Figure 55. Manhattan and QQ-plot of phenotype Diseases of pulp and periapical tissues. Figure 56. Manhattan and QQ-plot of phenotype Dental pulpitis. Figure 57. Manhattan and QQ-plot of phenotype Caries in clefts patients. Figure 58. Manhattan and QQ-plot of phenotype Cleft hard palate.

Figure 59. Manhattan and QQ-plot of phenotype Cleft lip, lip and palate or palate. Figure 60. Manhattan and QQ-plot of phenotype Cleft lip or cleft lip and palate. Figure 61. Manhattan and QQ-plot of phenotype Cleft lip. Figure 62. Manhattan and QQ-plot of phenotype Cleft palate. Figure 63. Manhattan and QQ-plot of phenotype Pathological resorption of teeth. Figure 64. Manhattan and QQ-plot of phenotype Diseases of salivary glands. Figure 65. Manhattan and QQ-plot of phenotype Stomatitis and related lesions. Figure 66. Manhattan and QQ-plot of phenotype Temporomandibular joint disorders. Figure 67. Manhattan and QQ-plot of phenotype Tooth wear.
